# Socio-economic inequalities and the COVID-19 epidemic in France: territorial analyses by epidemic wave and by metropolitan area

**DOI:** 10.1101/2025.05.20.25327911

**Authors:** Luka Canton, Pierre Schalkwijk, Jordi Landier, Stanislas Rebaudet, Emilie Mosnier, Pascal Handschumacher, Stève Nauleau, Philippe Malfait, Ludivine Launay, Cyrille Delpierre, Michelle Kelly Irving, Sabira Smaili, Stephanie Vandentorren, Jean Gaudart

## Abstract

**Background:** Previous studies highlighted the relationships between socioeconomic inequalities and the population’s risk to become disease or die during COVID-19 epidemic. In France, socioeconomic inequalities vary across metropolitan areas, but little is known if that could explain the spatial disparities observed in terms of incidence and testing rates. We assessed the impact of socioeconomic inequalities on testing and incidence rates of COVID-19 for each metropolitan area and wave.

**Methods:** For each of the 22 metropolitan areas, we relied on socioeconomic variables from census data to define socioeconomic profiles using classification on the principal components. We analysed associations between socioeconomic profiles, testing and incidence rates by epidemic wave from July-2020 to March-2023, using spatialised generalised additive mixed models. We performed meta-regressions to study the distribution of testing and incidence rate ratios (socially deprived vs privileged) across metropolitan areas, according to vaccination rates.

**Results:** Socially deprived metropolitan areas had lower testing rates than privileged, but during wave 4 (July-October-2021, extended health pass), testing rates increased in more deprived areas. Incidence rates were higher in deprived areas (waves 2-4, July-2020 to October-2021) but reversed between waves 6 to 9 (March-2022 to March-2023). Meta-regression analysis indicated that high vaccination coverage narrowed testing and incidence gaps between deprived and privileged.

**Conclusions:** The impact of social inequalities on the population’s testing and incidence COVID-19 epidemic was driven by socioeconomic inequalities across metropolitan areas and varied across epidemic waves. Vaccination rates and the presence of health measures (lockdowns, health pass) seem to help reduce these disparities.

## Background

Many studies have highlighted socioeconomic inequalities exacerbated during the COVID-19 pandemic, including in France, where people socially deprived had the highest severe impact, as demonstrated by the Epicov survey [1] and the Santé Publique France study [2]. Social health inequalities related to COVID-19 have been analysed at various geographical levels, from the departmental level, where inter-departmental disparities were observed [3–6], to the finer scale of IRIS (sub-municipal level) [7].

However, these studies were conducted within a global context that doesn’t always consider spatial heterogeneity and local geographical context, due to socio-economic and epidemic factors. Deprivation indices commonly used (French Deprivation Index (FDEP), European Deprivation Index (EDI)) to study social inequalities in health in France don’t always fully capture local socio-economic disparities. E.g., car ownership may not have the same significance among the EDI indicators in urban and rural areas [7,8].

Spatial variations in the spread of the COVID-19 epidemic varied according to the viral circulation but may also have resulted from the timing and the implementation of local protective measures. The test, trace, isolate strategy was implemented in France in May 2020 and was provided free of charge until October 2021. The first national lockdown was implemented between March and May 2020 and was restrictive, but the second (between October and December 2020) and the third (from April to May 2021) were less stringent. COVID-19 vaccination strategy was implemented in five stages from January 2021 onwards, with vaccination cost-free. Local restrictive measures could differ between areas: for example, a weekend lockdown was in place at the end of February 2021 in Alpes-Maritimes, then lockdowns were introduced in several departments in mid-March and extended to the rest of France between April and May. Specific interventions were also implemented, such as mobile teams of health mediators in deprived areas of Marseille [9]. Disparities in population demographics characteristics, the spatial and chronological variations in health protection measures across certain localities may have also contributed to epidemic variations within France.

A few ecological studies focusing on finer geographical areas (IRIS), such as the Provence-Alpes-Côte-d’Azur region [7,10], the Alpes-Maritimes department [11,12], and the municipalities of Paris [13], confirmed, using different methods, that the deprived population was most affected by the epidemic. No study analysed the epidemic dynamics of each metropolitan area to measure the differential impact on testing and incidence related to socio-economic inequalities in health.

Our study aimed to conduct a comparative analysis of testing and incidence rates across French metropolitan areas at the IRIS level, according to the socio-economic profiles of each metropolitan area. Our goal was to better understand the dynamics of the COVID-19 epidemic on socio-economic health inequalities across the 22 French metropolitan areas.

## Material and methods

### Study design

We conducted an ecological study of the COVID-19 epidemic in the 22 French metropolitan areas at a fine spatial scale, which are “grouped blocks for statistical information” (french acronym: IRIS, corresponding to an area comprising 2000 inhabitants relatively homogenous in terms of socioeconomic characteristics). In total, the 22 metropolitan areas included 7,756 IRIS.

### Study period

We analysed COVID-19 data on the number of tests and positive COVID-19 cases. We divided the data into several epidemic waves. These waves differed in terms of incidence and testing levels and health measures (lockdown, health pass, vaccination…). We defined waves in two steps.

- For waves 1 to 3, we used the dates determined by the French National Institute of Statistics and Economic Studies (INSEE) as presented in an illustration from a publication [15]. Since we did not have complete data for the first epidemic, we kept data from the start date of the second wave (6 July 2020).
- To identify the next waves, we examined the daily incidence rate in France, which was smoothed using a rolling mean 7-day. A new wave was considered to start when the incidence rate increased for at least 14 consecutive days and to end when it decreased for a minimum of 14 consecutive days. With this method, we identified five additional waves.

In total, we had 8 epidemic waves that defined our analysis periods (from 6 July 2020 to 4 March 2023) and we aggregated the number of tests and cases per wave.

### Variables

Data sources were described in the appendix (Supplementary Table S1).

### COVID-19 data

We used the number of tests and positive cases of COVID-19 on a rolling 7-day basis by IRIS from the French National Public Health Agency (Santé Publique France) SI-DEP information system, which collected and aggregated information about COVID-19 tests. We used France’s daily incidence rate from SI-DEP open access.

We used daily data to describe policy measures in France, provided by the Oxford COVID-19 Government Response Tracker [14]. We used an indicator between 0 and 100, where the higher the indicator, the more severe and numerous the measures.

We used weekly data on vaccination rates from the French health insurance at the scale of metropolitan areas.

### Socioeconomic data

We used socio-economic data by IRIS from INSEE to describe the population based on the national census of 2017 and the social and fiscal localised income (Filosofi). The selection of socioeconomic variables was made on the indicators and these dimensions (family and households; immigration and mobility; education; housing; and employment and income) to allow the creation of an indicator according to the method of Lalloué [15]. We used the 2017 European Deprivation Index (EDI) by IRIS, which we requested from MapInMed. Detailed information for each variable is provided in the appendix (Supplementary Table S2). These variables were used to build socioeconomic profiles by using hierarchical ascending clustering (HAC) on principal components applied to each metropolitan area [3]. Maps of socioeconomic profiles are available in the appendix (Supplementary Figures S1– S22).

### IRIS selection

First, we excluded activity and diverse IRIS to keep only residential IRIS. Second, we deleted IRIS with fewer than 30 households, as many variables focused on household and housing characteristics, which may be unreliable in such small units. IRIS with missing data on the median income were excluded, except if it was available at the scale of the municipality. We deleted IRIS with a higher number of positive cases than the number of inhabitants on several 7-day rolling periods, likely due to the misallocation of test results. Finally, a deprived IRIS in the Nice metropolitan area with a high testing rate was eliminated because of the leverage effect it exerted in the multivariate testing models, which risked skewing results by overrepresenting an atypical dynamic.

### Statistical analysis

Statistical analyses, as graphical and cartographic representations, were performed using R software (version 4.2.2., R Development Core Team, R Foundation for Statistical Computing, Vienna, Austria).

### Trend analysis

For each epidemic wave, we reported the median EDI of the most deprived population in each metropolitan area and examined its relationship with incidence and testing rates across the 22 metropolitan areas and 8 waves. We used LOESS regression curves, with 95% confidence intervals, to capture trends between deprivation with testing and incidence, allowing the identification of deprived areas that deviated from general patterns. To assess how incidence and testing for each metropolitan area evolved, we classified them during each wave according to their position in relation to the LOESS curve and its confidence intervals (i.e., below the CI, within the CI and below the curve, within the CI and above the curve, and above the CI). These categories were tracked over waves using heatmaps and Sankey diagrams, allowing temporal changes to be visualised and persistent or evolving patterns in epidemic dynamics to be identified (Supplementary Figures S23–S24). All these graphs were performed on positivity rates (Supplementary Figure S25).

Finally, graphs were produced to compare cumulative median vaccination rates per wave (at the scale of metropolitan area) with the median EDI and testing rates of the most deprived IRIS, to assess whether social inequalities in testing were also visible in vaccination levels.

### Statistical model

We used generalised additive mixed models (GAMMs) to quantify the difference in testing rate and incidence rate between the least and most deprived profiles. We ran one GAMM for each wave and each metropolitan area. We used a negative binomial distribution to account for the overdispersion of the data, as appropriate for counts and we included the logarithm of the population as an offset. The main variable of interest was the socio-economic profiles and the reference modality was the least socially deprived profile. We adjusted on: the healthcare variables (localised potential accessibility at the municipality scale (LPA), number of pharmacies and medical laboratories, number of retirement homes), the logarithm of population density, the population’s age distribution and environmental profiles (refer to the appendix for more detailed information in the « adjustment variables » section). Incidence rate models included the logarithm of the number of tests per person per IRIS as an adjustment variable. We included a random effect at the municipality level, taking into account similarities for IRIS in the same municipality. We used a random Markov field, which allowed us to account for spatial autocorrelation of IRIS based on the Queen’s criterion of contiguity. With the results, we mapped adjusted testing and incidence rate ratios (aTRRs, aIRRs) for the most deprived socioeconomic profile. We ran univariate models (only with random effect and spatial autocorrelation as factors) and mapped testing and incidence rate ratios (TRRs, IRRs) (Supplementary Figure S26). For the Nice metropolitan area, univariate and multivariate models on the testing rates with and without the IRIS outlier were run (Supplementary Table S3).

Model on the testing rates (including spatial autocorrelation and random effect) :

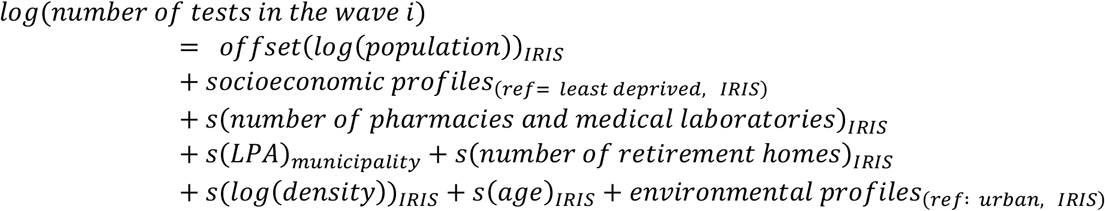

Model on the incidence rate (including spatial autocorrelation and random effect) :

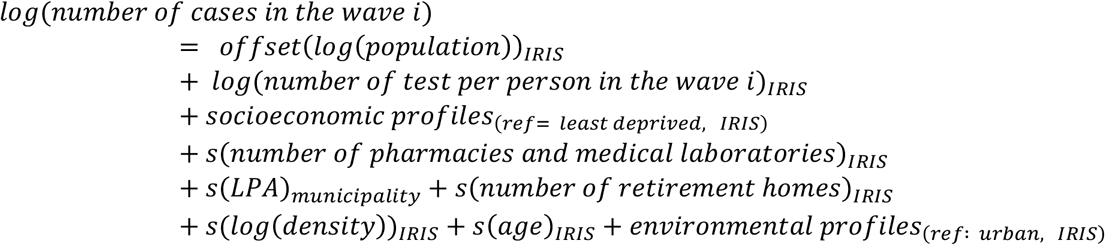

### Meta-regression on aTRRs and aIRRs of the deprived population

We used results of the previous models in linear meta-regressions to explain aTRRs and aIRRs, which respectively represent the differences in testing and incidence rates between the least deprived and most privileged profiles within each metropolitan area. We included three covariates as linear predictors: the number of waves, the median cumulative vaccination rate of the wave in each metropolitan area, and the median EDI of the most deprived people in each metropolitan area. To help with interpretation, we plotted the mean of aTRRs and aIRRs per wave. As sensitive analysis, we ran linear meta-regressions on TRRs and IRRs provided by univariate models (Supplementary Figures S27– S28).

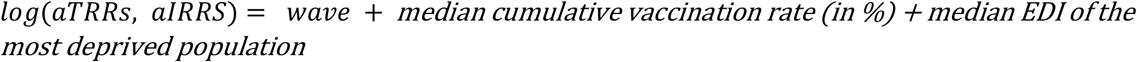

## Results

### IRIS selection

Of the 7756 IRIS, we kept 7186 for analysis. Exclusions included 554 non-residential IRIS, 6 with fewer than 30 households, 8 with missing data on median income and due to consistently higher COVID-19 cases compared to the population over several weeks (Table 1). In Nantes, one IRIS had more cases than inhabitants on several 7-day rolling periods during wave 5 (rolling weeks from 12 January 2022 to 29 January 2022). One deprived IRIS in Nice was deleted, which has a leverage effect on testing rates in the multivariate models due to its particularly high testing rate (Table 1). Exceptionally, in models, due to missing data on testing and incidence for certain IRIS for waves 7 to 9, these IRIS were excluded from the models only for these periods. These include 1 IRIS in Toulouse, 3 in Grand Paris and 3 in Lyon.

**Table 1:**
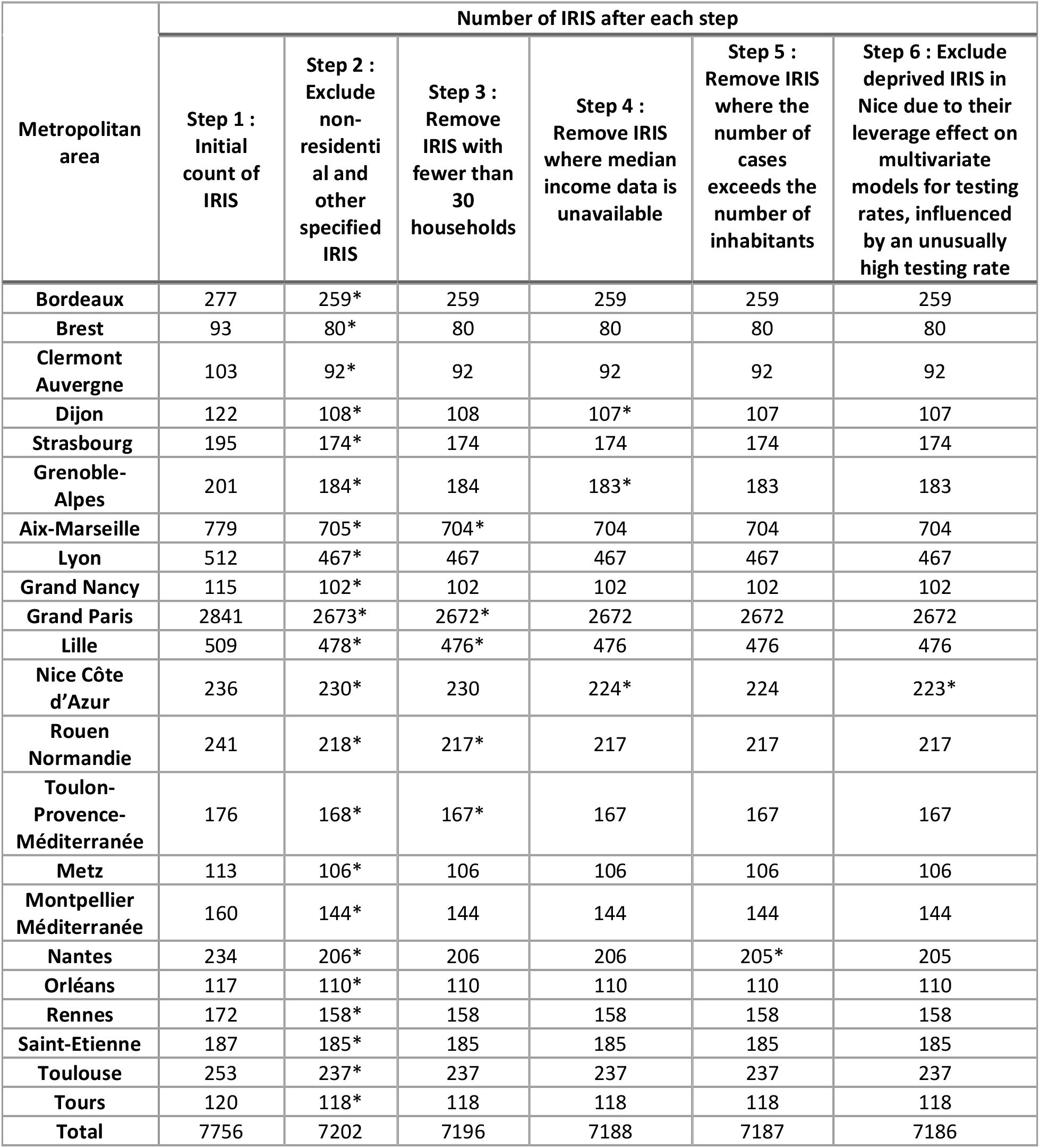
Number of IRIS for each French metropolitan area after each selection step. Numbers with * indicate a decrease compared to the previous step.

| Metropolitan area | Number of IRIS after each step |  |  |  |  |  |
| --- | --- | --- | --- | --- | --- | --- |
|  | Step 1 : Initial count of IRIS | Step 2 : Exclude non-residential and other specified IRIS | Step 3 : Remove IRIS with fewer than 30 households | Step 4 : Remove IRIS where median income data is unavailable | Step 5 : Remove IRIS where the number of cases exceeds the number of inhabitants | Step 6 : Exclude deprived IRIS in Nice due to their leverage effect on multivariate models for testing rates, influenced by an unusually high testing rate |
| Bordeaux | 277 | 259* | 259 | 259 | 259 | 259 |
| Brest | 93 | 80* | 80 | 80 | 80 | 80 |
| Clermont Auvergne | 103 | 92* | 92 | 92 | 92 | 92 |
| Dijon | 122 | 108* | 108 | 107* | 107 | 107 |
| Strasbourg | 195 | 174* | 174 | 174 | 174 | 174 |
| Grenoble-Alpes | 201 | 184* | 184 | 183* | 183 | 183 |
| Aix-Marseille | 779 | 705* | 704* | 704 | 704 | 704 |
| Lyon | 512 | 467* | 467 | 467 | 467 | 467 |
| Grand Nancy | 115 | 102* | 102 | 102 | 102 | 102 |
| Grand Paris | 2841 | 2673* | 2672* | 2672 | 2672 | 2672 |
| Lille | 509 | 478* | 476* | 476 | 476 | 476 |
| Nice Côte d'Azur | 236 | 230* | 230 | 224* | 224 | 223* |
| Rouen Normandie | 241 | 218* | 217* | 217 | 217 | 217 |
| Toulon-Provence-Méditerranée | 176 | 168* | 167* | 167 | 167 | 167 |
| Metz | 113 | 106* | 106 | 106 | 106 | 106 |
| Montpellier Méditerranée | 160 | 144* | 144 | 144 | 144 | 144 |
| Nantes | 234 | 206* | 206 | 206 | 205* | 205 |
| Orléans | 117 | 110* | 110 | 110 | 110 | 110 |
| Rennes | 172 | 158* | 158 | 158 | 158 | 158 |
| Saint-Etienne | 187 | 185* | 185 | 185 | 185 | 185 |
| Toulouse | 253 | 237* | 237 | 237 | 237 | 237 |
| Tours | 120 | 118* | 118 | 118 | 118 | 118 |
| Total | 7756 | 7202 | 7196 | 7188 | 7187 | 7186 |

### Description of waves

The temporal limits for each wave could be visualised on the two graphs showing the incidence rate and the index describing the number of protective measures in the degree of restriction (Figure 1). We observed different amplitudes of the epidemic waves, with waves 5 to 9 having higher incidence rates than the first periods (Figure 1A).

**Figure 1:**
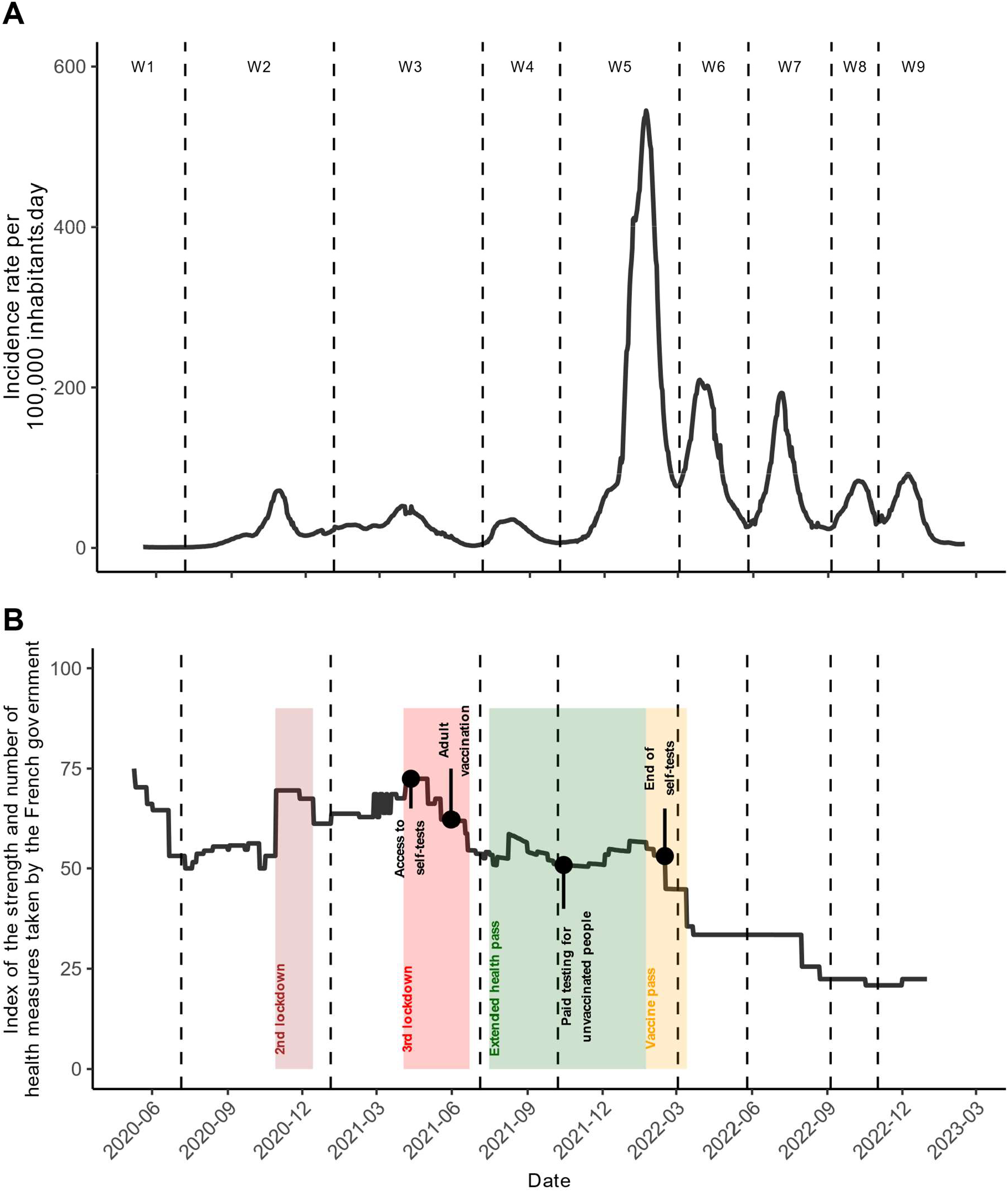
Daily evolution of the incidence rate of COVID-19 (A) and the index (varying from 0 to 100) of the accumulation and rigour of the government’s health measures according to the Oxford index (B), France, May 2020 - February 2023. In figure A, the curve represents a 7-day moving average of incidence rates. The dotted lines correspond to the boundaries of the epidemic waves from wave 2 (W2) to wave 9 (W9). Data sources: SI-DEP ; Blavatnik School of Government, Université d’Oxford

A higher index characterised waves 1 to 3 due to lockdown and curfews (Figure 1B). The beginning of the 3rd wave was the start of the vaccination for people aged over 75 (January 2021) and was extended for people aged over 18 later (May 2021). Self-tests were introduced in this 3rd wave (Figure 1B). Then, the index decreased during waves 4 and 5 with the presence of a health pass, which was later converted into a vaccinal pass (Figure 1B). The beginning of wave 5 corresponded to tests for which a fee was charged to those not vaccinated (Figure 1B). We observed a fall in the index for the last waves with fewer restrictive health measures (Figure 1B).

#### Testing and incidence rates among deprived profiles for each metropolitan area

We focused on the subset of IRIS identified as most deprived in the clustering process at the metropolitan area level. For each metropolitan area, we computed the median deprivation index (EDI) for the most deprived subset.

During wave 4, the most deprived with a high median EDI had elevated testing rates, especially at Aix-Marseille (AMP, > 9.000 tests/100.000 inhabitants.week) (Figure 2A). We observed the same trend during waves 5 and 7 (Figure 2A). Heatmaps showed that some deprived areas from metropolitan areas had high testing rates (Strasbourg (STR), Aix-Marseille (AMP), Lyon (LYN)) or low (Toulon (TLN), Orléans (ORL), Montpellier (MTP) and Clermont (CLT)) for at least 4 waves than expected given their median EDI (Supplementary Figure S23).

**Figure 2:**
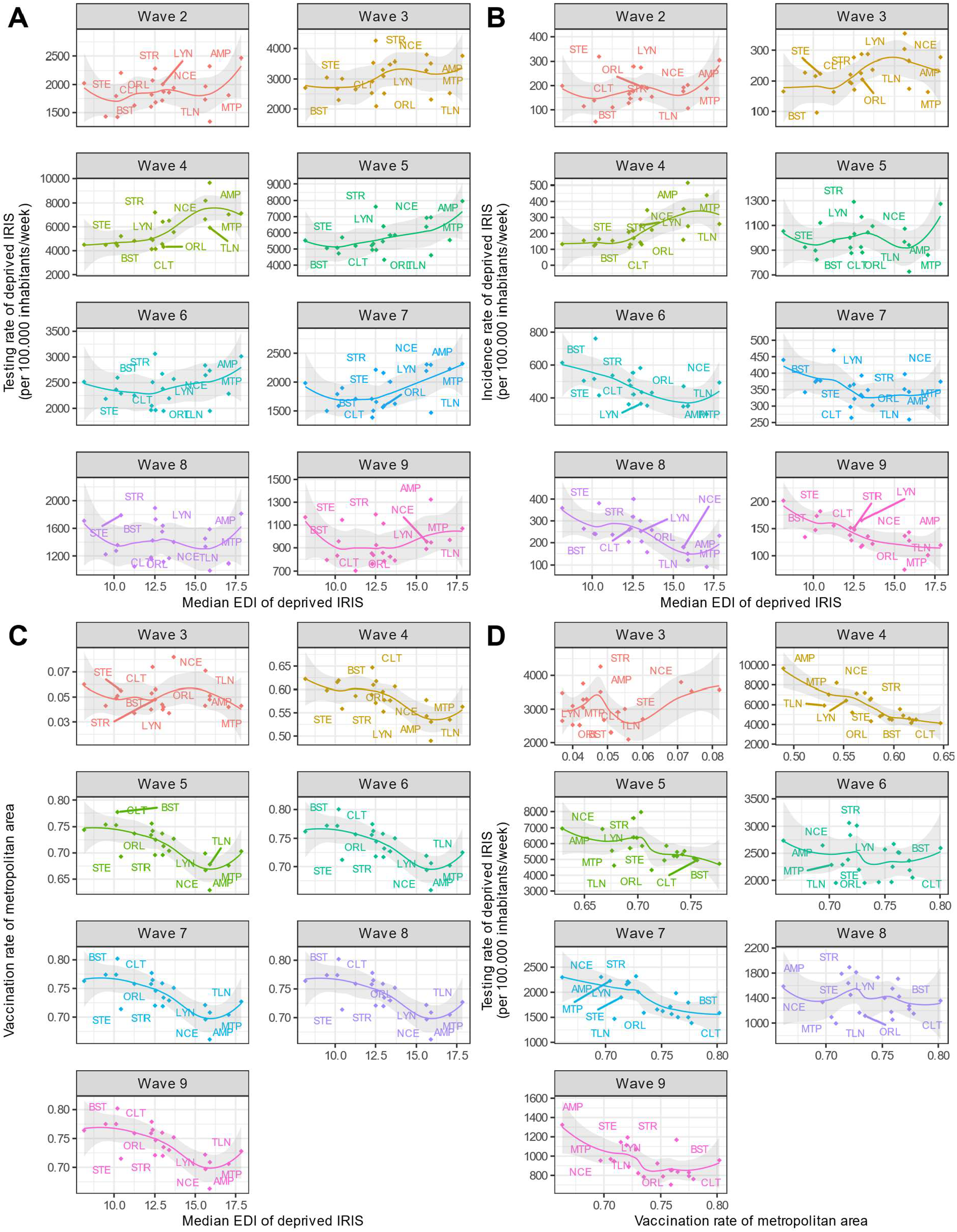
Evolution by wave (waves 2 to 9) of testing rates (A) and incidence rates (B) for IRIS belonging to the most deprived profile, median vaccination rates of metropolitan areas (C) in relation to the median deprivation score (EDI) for IRIS belonging to the most deprived profile in the 22 French metropolitan areas, and testing rates compared to the median vaccination rate per metropolitan area (D), France, July 2020 – March 2023, n=1096 IRIS deprived, n=22 french metropolitan areas. AMP = Aix-Marseille-Provence ; BST = Brest ; CLT = Clermont Auvergne ; LYN = Lyon ; MTP = Montpellier Méditerranée ; NCE = Nice Côte d’Azur ; ORL = Orléans ; STE = Saint-Étienne ; STR = Strasbourg ; TLN = Toulon ; EDI: European deprivation index Testing rates, incidence rates, and median EDI scores were computed only for IRIS belonging to the most deprived profile of each metropolitan area. Vaccination data were available at the metropolitan area level. For each wave, the median cumulative vaccination rate corresponded to the cumulative rate at the midpoint of the wave. The four graphs showed smoothed curves (LOESS regressions) with 95% confidence intervals shaded in grey. Data sources: SIDEP-Santé publique France, MapInMed, French health insurance

Incidence rates were higher in metropolitan areas, with a median EDI elevated among deprived populations during waves 3 and 4 (Figure 2B). Since wave 6, the trend reversed, and metropolitan areas with the highest EDI had the lowest incidence rate (Figure 2B). Positivity rates followed the same trend (Supplementary Figure S25). Heatmaps revealed persistently high incidence rates over 4 four waves in some metropolitan areas (Nice (NCE), Lyon (LYN), Saint-Étienne (STE), and Strasbourg (STR)) while others showed consistently lower rates (Montpellier (MTP), Toulon (TLN), and Orléans (ORL)) than expected given their median EDI (Supplementary Figure S24).

Low vaccination rates (at the scale of metropolitan area) corresponded to low median EDI of deprived areas, except for wave 3 (Figure 2C). Aix-Marseille (AMP) had the lowest vaccination rates (< 65%), while Brest (BST) had the highest rates (> 80%) (Figure 2C). This relationship between deprivation and vaccination rates was observed with testing rates (Figure 2D). Especially during wave 4, metropolitan areas with low vaccination rates had higher testing rates (Figure 2D). A similar trend was observed during waves 5, 7 and 9 (Figure 2D).

### Comparison of differences in testing and incidence rate between the most deprived and most privileged profiles in each metropolitan area, by wave

Testing rates were lower in the deprived population (Figure 3A). However, differences in aTRRs were greater after wave 4, especially for waves 8 and 9 for some metropolitan areas (E.g. Toulouse, wave 2: aTRR = 0.83, 95%IC = [0.71 0.97]; wave 9: aTRR = 0.62, [0.51 0.75]) (Figure 3A). At wave 4, no significant differences existed between the aTRRs for 18 metropolitan areas, which were all close to 1 (Figure 3A). From wave 5 onwards (Omicron variant, introduction of fee-based tests for unvaccinated people), disparities between metropolitan areas increased. For example, in wave 8, Toulon had an aTRR of 0.60 [0.44–0.82], while Grand Paris reached 0.85 [0.81–0.88] (Figure 3A). The same general interpretation could be made with TRRs (Supplementary Figure S26A).

**Figure 3:**
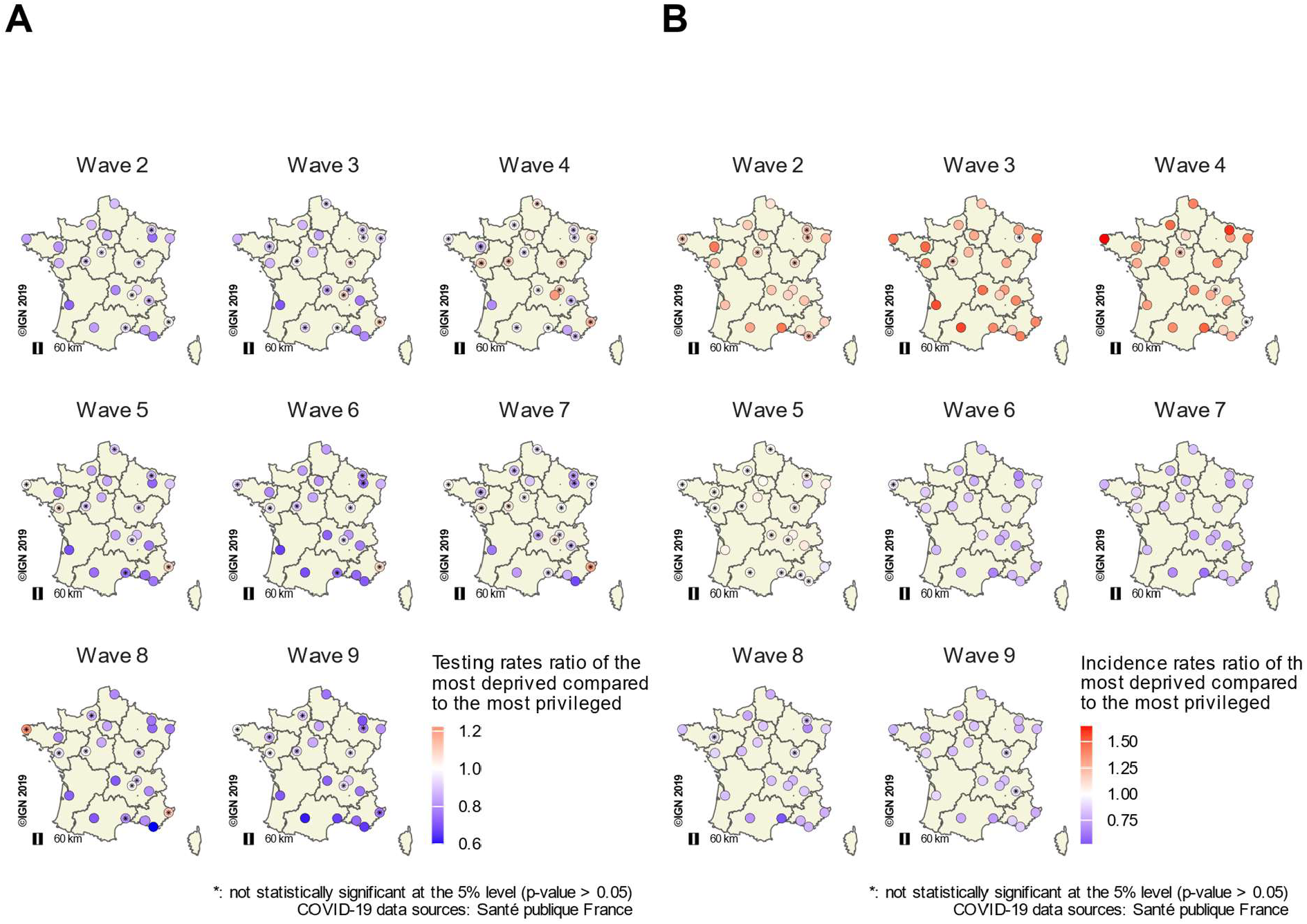
Spatial distribution of testing (A) and incidence (B) rate ratios between the most deprived and most privileged profiles in the 22 French metropolitan areas, by wave (waves 2 to 9), France, July 2020 – March 2023, n=7186 IRIS, n=22 French metropolitan areas. *: not statistically significant at the 5% level (p-value > 0.05) The ratios were computed from multivariate GAM models per wave, comparing testing and incidence rates for each metropolitan area between the socioeconomic profile with the lowest average EDI (the least deprived) and the other profiles. Data sources: SIDEP-Santé publique France

Incidence results could be summarised in three phases. Firstly, from waves 2 to 4, aIRRs were greater than 1 for all metropolitan areas, indicating that incidences were highest in the deprived population (Figure 3B). Secondly, during wave 5 (spread of the Omicron variant), aIRRs were close to 1 for most metropolitan areas (Figure 3B). Lastly, from waves 6 to 9, the trend reversed: incidences were lower in deprived compared to privileged areas (aIRRs < 1) (Figure 3B). We observed the same trends with IRRs (Supplementary Figure S26B).

### Meta-regression on adjusted testing rate ratios and incidence rate ratios of deprived profiles

On average, aTRRs were lower than 0.92 (deprived areas under-tested) for each wave except for wave 4 (mean aTRRs = 1) (Figure 4A). Generally, aTRRs decreased along waves, but increased between wave 3 (mean aTRRs = 0.9) and 4 and between waves 6 (mean aTRRs = 0.82) and 7 (mean aTRRs = 0.92) (Figure 4A).

**Figure 4:**
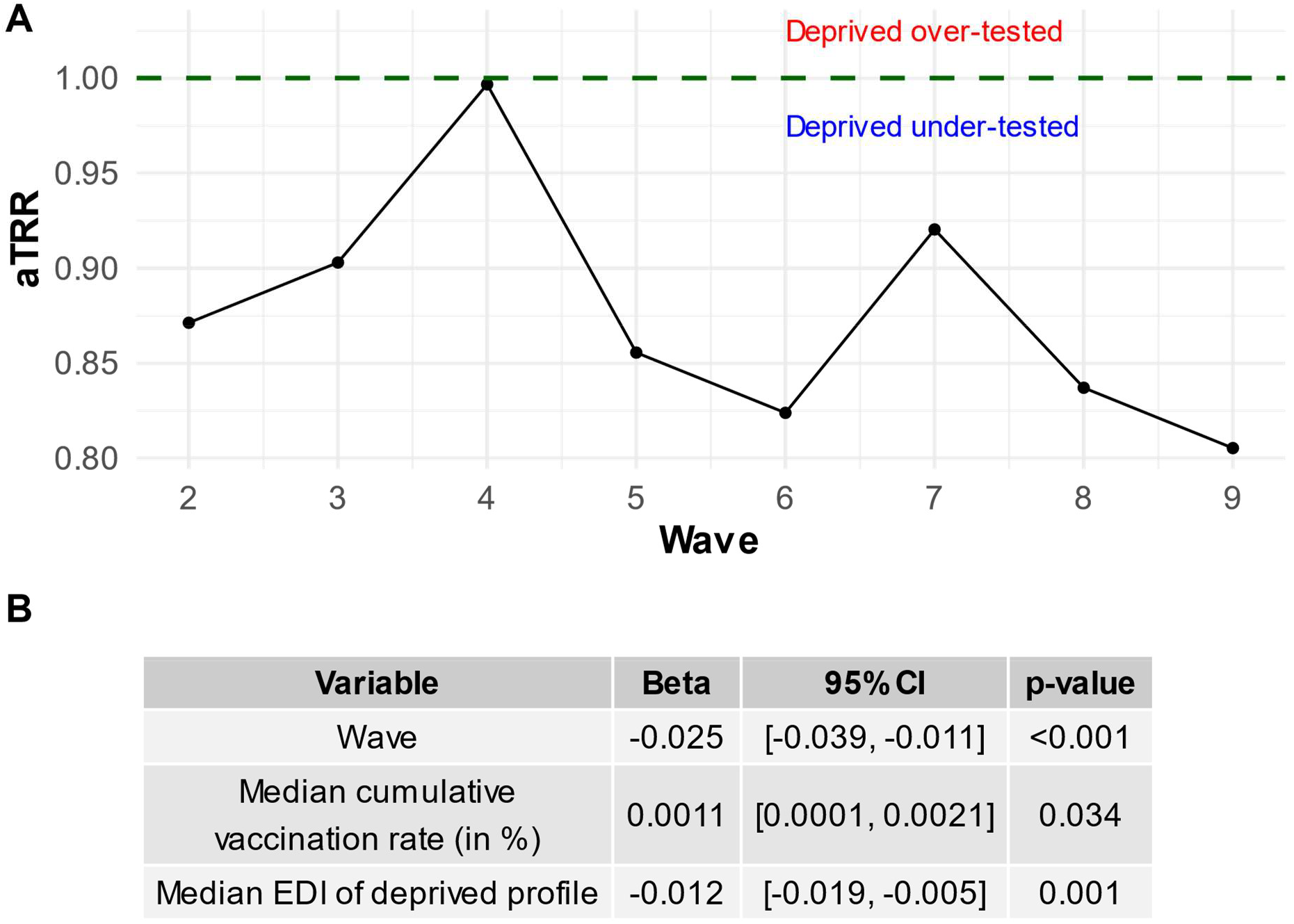
Mean of aTRRs between the most deprived and most privileged profiles of the 22 French metropolitan areas per epidemic wave (A) and results of meta-regression on logarithm of aTRRs (B), France, July 2020 – March 2023, n=176. aTRR : adjusted testing rate ratio, CI: confidence intervals, EDI: European deprivation index Green dotted lines corresponded to the testing equality point between the privileged and the deprived (aTRR = 1). The table showed estimated betas for the three explanatory variables (wave, median cumulative vaccination rate and median EDI of deprived profile) with confidence intervals (95% CI) and p-values. Data sources: SIDEP-Santé publique France, MapInMed, French health insurance

Meta-regressions confirmed that aTRRs significantly decreased with successive waves (β = –0.025), suggesting a progressive worsening of under-testing in deprived areas (Figure 4B). Higher cumulative vaccination rates were linked to higher aTRRs (β = 0.0011) (Figure 4A). Under-testing of deprived areas became more balanced (if aTRRs < 1 at low vaccination rates), or their over-testing intensified (if aTRRs > 1 at low vaccination rates) (Figure 4A). A higher median EDI was linked to lower aTRRs (β = –0.012) (Figure 4A). Over-testing of deprived areas receded gradually (if aTRRs > 1 at low EDI) or worsened their under-testing (if aTRRs < 1 at low EDI) (Figure 4A). Meta-regressions on TRRs presented the same effects of the 3 variables (Supplementary Figure S27A-B).

On average, aIRRs were higher than 1.2 (deprived areas in over-incidence) during waves 2 to 4 and lower than 0.82 (deprived areas in under-incidence) between waves 6 and 9 (Figure 5A). During wave 5, aIRRs were close to 1 (mean aIRRs = 1.02) (Figure 5A).

**Figure 5:**
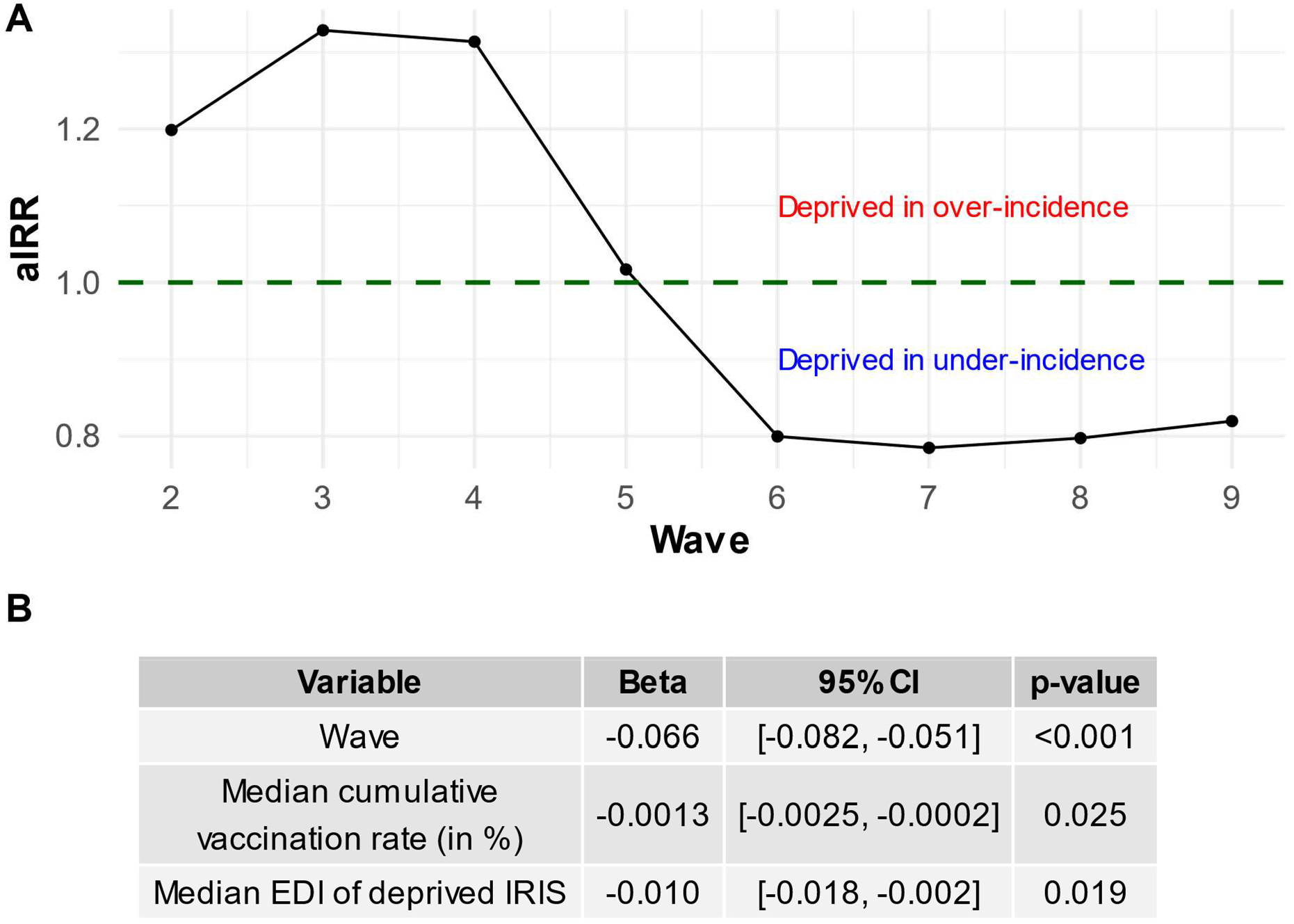
Mean of aTRRs between the most deprived and most privileged profiles of the 22 metropolitan areas per epidemic wave (A) and results of meta-regression on logarithm of aTRRs (B), July 2020 – March 2023, (n=176). aIRR: adjusted incidence rate ratio, CI: confidence intervals, EDI: European deprivation index Green dotted lines corresponded to the testing equality point between the privileged and the deprived (aIRR = 1). Results showed estimated betas for the three explanatory variables (wave, median cumulative vaccination rate and median EDI of deprived profile) with confidence intervals (95% CI) and p-values. Data sources: SIDEP-Santé publique France, MapInMed, French health insurance

Meta-regressions confirmed that waves were linked to a significant decrease in aIRRs (β = –0.07) (Figure 5B). Higher cumulative vaccination rates were also linked to lower aIRRs (β = -0.0013) (Figure 5B). Over-incidence of deprived areas decreased (if aTRRs > 1 at low vaccination rates) or their under-incidence worsened (if aIRRs < 1 at low vaccination rates) (Figure 5B). A higher median EDI was associated with lower aIRRs (β = –0.01) (Figure 5B). Over-incidence of deprived areas decreased (if aIRRs > 1 at low EDI) or their under-incidence worsened (if aIRRs < 1 at low EDI) (Figure 5B). Meta-regressions on IRRs presented the same results for the effect of waves (Supplementary Figure S28A-B). There were no statistically significant effects for the median cumulative vaccination rate and median EDI (p-values < 0.16), but the signs of the coefficients remained consistent (Supplementary Figure S28B).

## Discussion

This study assessed the impact of socioeconomic inequalities on COVID-19 testing and incidence across metropolitan areas in mainland France. By constructing local socio-economic profiles, we gained insights into the specific contexts of each metropolitan area. We used EDI to identify the most deprived profile in each metropolitan area and showed that deprived profiles could display substantial differences in EDI between metropolitan areas.

Testing and incidence rates fluctuated over time among deprived areas. During wave 4, testing increased in more deprived areas, while it declined in highly vaccinated metropolitan areas. From wave 6 onwards, incidence rates dropped in deprived IRIS with lower EDI. However, heatmaps and Sankey diagrams suggested that EDI alone could not fully explain these patterns, highlighting the need for localised analysis.

We showed that deprived populations were generally less tested, except in wave 4, when testing rates were more homogenous between deprived and privileged profiles. This may be related to implementing the health pass, which required vaccination or a recent negative test for access to public places and work, especially for frontline workers, who belong to a more deprived population. This obligation may have temporarily increased testing in these populations during wave 4. Meta-regression suggested that high vaccination rates of metropolitan areas reduced under-testing of deprived areas, as we observed during wave 4. These differences may be explained by a higher vaccination rate among the privileged, who had better access to vaccination [16–18]. From wave 5 onward, testing became payment for unvaccinated people, contributing to a decline in testing among the most deprived, for whom the cost may have been a barrier [19,20]. This was consistent with our finding that testing gaps widened over time. Finally, the largest gaps in testing were observed in metropolitan areas where the median EDI of the deprived was highest according to the meta-analysis.

Regarding incidence, deprived populations were more affected during waves 2 to 4. From wave 6 onwards, the trend reversed, with privileged IRIS showing higher incidence rates. As vaccination was higher among the privileged population [16–18], this may also be explained by greater protection for the privileged during waves 3 and 4. However, vaccination may have a reduced protective effect against the Omicron variant (since wave 5) [21], which was more contagious than its predecessors [22]. Also, we hypothesised that privileged populations enjoyed stronger protection when stringent health measures were applied. Once these restrictions eased from wave 6 onward, their social habits, such as more frequent mobility, leisure activities, and social gatherings, may have increased exposure. Meta-analysis showed that high vaccination rates could correct the over-incidence of the deprived. It highlighted a potential reduction of the over-incidence of the deprived when their median EDI rose, or increased differences in incidence in favour of the deprived. We observed that on the graph, incidence and positivity rates declined in deprived IRIS with lower EDI since wave 6. The decrease in restrictive measures since wave 6, combined with the spread of the Omicron variant, could explain why the incidence was lower in the most deprived populations.

Several study limitations must be acknowledged. Statistical models may have been affected by multicollinearity between socio-economic variables and population density. Nonetheless, observed trends remained consistent across different metropolitan areas and waves and according to sensitive analysis. There may also be biases linked to the structure of metropolitan areas as Nice, where differences were lower between profiles may be due to the presence of mountainous and rural areas to the north of the area, where access to testing is more difficult. This point had already been raised in a previous study of the PACA region [7]. This underlines the importance of the choice of the spatial scale and the study area, which could affect results in measuring social inequalities in health, and of carrying out analyses on urban areas.

Another limitation was the absence of data on vaccination at the IRIS level, which could have provided additional information on incidence and testing behaviour. However, the hesitancy of some people to be tested, apart from the cost of the tests, could also be due to low health literacy, less confidence in the healthcare system and a fear of stigmatisation in the event of a positive test [7,19,24]. Addressing the behavioural domain is complex, due to the study design, where individual determinants cannot be taken into account. However, using mobility data would also provide a better picture of these behaviours during periods when health measures were in application [25,26].

Finally, this study was confronted with ecological bias, where results cannot necessarily be applied on an individual scale [3,27]. However, as social determinants in individual data remain sparse in France, studying territorial inequalities in each of these metropolitan areas on a fine geographical scale reduced this bias by better capturing the local socioeconomic contexts and their epidemic dynamics. This approach allowed for more precisely targeted public health interventions.

## Conclusion

In conclusion, this study highlighted the significant role of socioeconomic inequalities in COVID-19 testing and incidence across metropolitan areas in mainland France. Testing and incidence gaps between privileged and deprived populations were linked not only to the level of deprivation of the most deprived profile of metropolitan areas, but were also associated with the protective measures as vaccination rates, lockdown and health pass. The study underlined the need for localised analysis and targeted public health interventions to address health inequalities.

## Ethical statement

This study was approved by the Ethics Committee of Aix-Marseille University (number 2022-10-20-006).

## Acknowledgments

We would like to thank Santé publique France, with which we collaborated on the Van Gogh project and which provided us with the COVID-19 data. Our sincere thanks also go to the Ligue Nationale Contre le Cancer, which provided us with the European Deprivation Index (EDI). We thank Mediate4Health and Prospective Cooperation for their collaborative work.

## Data availability

The aggregated data are accessible to researchers upon reasonable request for data sharing to the corresponding author. Requests for data require approval by Santé Publique France. Requests to access these datasets should be directed to the corresponding author.

## Authors’ contributions

JG, JL and SR designed the study. JG coordinated the project. LC, PS and JG did the data management and analysed data. LC wrote the first draft. LC, PS, JL, SV, SS, CD, and JG interpreted the results, with the help of SR, EL, EM, PH, FAR, FF ; SN, LL, PM, CD, MKI, SS. All authors reviewed and approved the final version of the manuscript.

