## Supplementary material for "Socio-economic inequalities and the COVID-19 epidemic in France: territorial analyses by epidemic wave and by metropolitan area"

### Appendix

#### Data sources

*Table S1: Data sources*

| Database name | Source | URL |
|---|---|---|
| Census population of 2017 (IRIS scale) | INSEE | https://www.insee.fr/fr/information/4467366 |
| Median income from Filosofi database (2018) | INSEE | https://www.insee.fr/fr/statistiques/5055909 (IRIS scale)<br>https://www.insee.fr/fr/statistiques/5009218 (municipality scale) |
| European deprivation index (EDI)(2017) | MapinMed | https://anticipe.eu/plateformes/MAPinMED |
| Equipment database (2021) | INSEE | https://drees2-sgsocialgouv.opendatasoft.com/explore/dataset/530_l-accessibilite-potentielle-localisee-apl/information/. |
| Localised potential accessibility (LPA)(2021) | DREES | https://drees2-sgsocialgouv.opendatasoft.com/explore/dataset/530_l-accessibilite-potentielle-localisee-apl/information/. |
| Land cover (2018) | Copernicus | https://land.copernicus.eu/pan-european/corine-land-cover/clc2018?tab=download. |
| Index of government stringency (2020-2023) | Blavatnik School of Government, Université d'Oxford | https://github.com/OxCGRT/covid-policy-tracker/tree/master/data. |
| Vaccination COVID-19 (2021-2023) | French health insurance | https://datavaccin-covid.ameli.fr/explore/dataset/donnees-de-vaccination-par-epci/information/. |
| Incidence rates for Mainland France (2020-2023) | SI-DEP | https://www.data.gouv.fr/fr/datasets/donnees-de-laboratoires-pour-le-depistage-a-compter-du-18-05-2022-si-dep/. |
| Shape of 200m tiles (to compute population density)(2017) | INSEE | https://www.insee.fr/fr/statistiques/6215138?sommaire=6215217 |
| Shape of IRIS (2019) | Geoservices | https://geoservices.ign.fr/contoursiris |

#### Socioeconomic variables and maps

*Table S2: Description of socioeconomic indicators used to create socioeconomic profiles*

| Libellé de la variable | **Nom de la variable** |
|---|---|
| Proportion of inhabitants in single-person households among the total household population | FM_men_seul |
| Average household size | FM_taille_men |
| Proportion of inhabitants in single-parent families among the total household population | FM_fam_mono |
| Proportion of inhabitants in couples without children in the total household population | FM_couple_sans_enf |
| Proportion of inhabitants in couples with children in the total household population | FM_couple_avec_enf |
| Proportion of married or in civil union in the population | FM_pacse_mariage |
| Proportion of inhabitants outside of households in the total population | FM_hors_men |
| Proportion of foreign inhabitants in the total population | IM_etrangers |
| Proportion of immigrants in the total population | IM_immigrés |
| Proportion of inhabitants in households that moved in less than 4 years ago | IM_emménage_moins_4ans |
| Proportion of inhabitants in households that moved in more than 10 years ago | IM_emménage_plus_10ans |

| | |
|---|---|
| Proportion of children aged 2 to 17 enrolled in school among the population of 2 to 17-year-olds | ED_2a17ans_scol |
| Proportion of students aged 15 to 64 in the population of 15-64-year-olds | ED_15ans_etplus_scol |
| Proportion of inhabitants without a diploma or holding a certificate among those over 15 years old (not in school) | ED_sansDip |
| Proportion of inhabitants with a CAP, BEP among those over 15 years old (not in school) | ED_CAP_BEP |
| Proportion of inhabitants with a baccalaureate, professional certificate among those over 15 years old (not in school) | ED_Bac |
| Proportion of inhabitants with a higher education diploma (at least a 2-year degree) among those over 15 years old (not in school) | ED_DipSup |
| Average number of rooms in main residences | LOG_pieces_res |
| Proportion of main residences built before 1970 | LOG_res_av1970 |
| Proportion of main residences built from 1991 (maximum 2017) | LOG_res_apr1990 |
| Proportion of main residences under 40m² | LOG_res_moins40m |
| Proportion of main residences over 100m² | LOG_res_plus100m |
| Proportion of main residences without a bathtub or shower | LOG_res_sans_douche |
| Proportion of main residences without central heating or individual electric heating | LOG_res_sans_chauffage |
| Proportion of households with a parking space | LOG_men_stationnement |
| Proportion of households without a car | LOG_men_sans_voiture |
| Proportion of households with two or more cars | LOG_men_voitures_2plus |
| Proportion of main residences (excluding studios) with 1 person in overcrowding | LOG_res_surrocup |
| Proportion of secondary residences and occasional housing among the total housing stock | LOG_res_secondaires |
| Proportion of people living in empty low-rent housing main residences among the total population | LOG_habitants_hlm_loué |
| Average number of rooms in main residences | LOG_nombre_pieces_personnes |
| Proportion of houses in housing stock | LOG_maisons |
| Proportion of apartments in housing stock | LOG_apparts |
| Proportion of people who don't own their main residence | LOG_res_non_proprio |
| Proportion of active men in the male population | ER_hommes_actifs |
| Proportion of active women in the female population | ER_femmes_actives |
| Proportion of active inhabitants in the total population | ER_pop_actifs |
| Proportion of unemployed inhabitants aged 15-24 among the active population of 15-24 years old | ER_chom_jeunes |
| Proportion of unemployed inhabitants aged 55-64 among the population of 55-64 years old | ER_chom_agé |
| Proportion of unemployed inhabitants in the active population | ER_chom |
| Proportion of farmers in the population aged 15 and older | ER_agric |
| Proportion of craftsmen in the population aged 15 and older | ER_artisan |
| Proportion of white-collar workers in the population aged 15 and older | ER_cadres |
| Proportion of intermediate professions in the population aged 15 and older | ER_profession_inter |
| Proportion of employees in the population aged 15 and older | ER_employe |
| Proportion of blue-collar workers in the population aged 15 and older | ER_ouvriers |
| Proportion of retirees in the population aged 15 and older | ER_retraites |
| Proportion of inhabitants without professional activity in the population aged 15 and older | ER_sans_activite_pro |
| Proportion of part-time workers in the population aged 15 and older | ER_temps_partiel |
| Proportion of self-employed inhabitants in the population aged 15 and older | ER_non_salarie |
| Proportion of inhabitants on permanent contracts (or in the public sector) in the active population | ER_cdi |
| Proportion of unstable employment in the active population (internships, fixed-term contracts, aid jobs, temporary work, apprenticeships) | ER_emploi_instable |
| Median income | ER_revenu_median |
| European deprivation index | EDI |

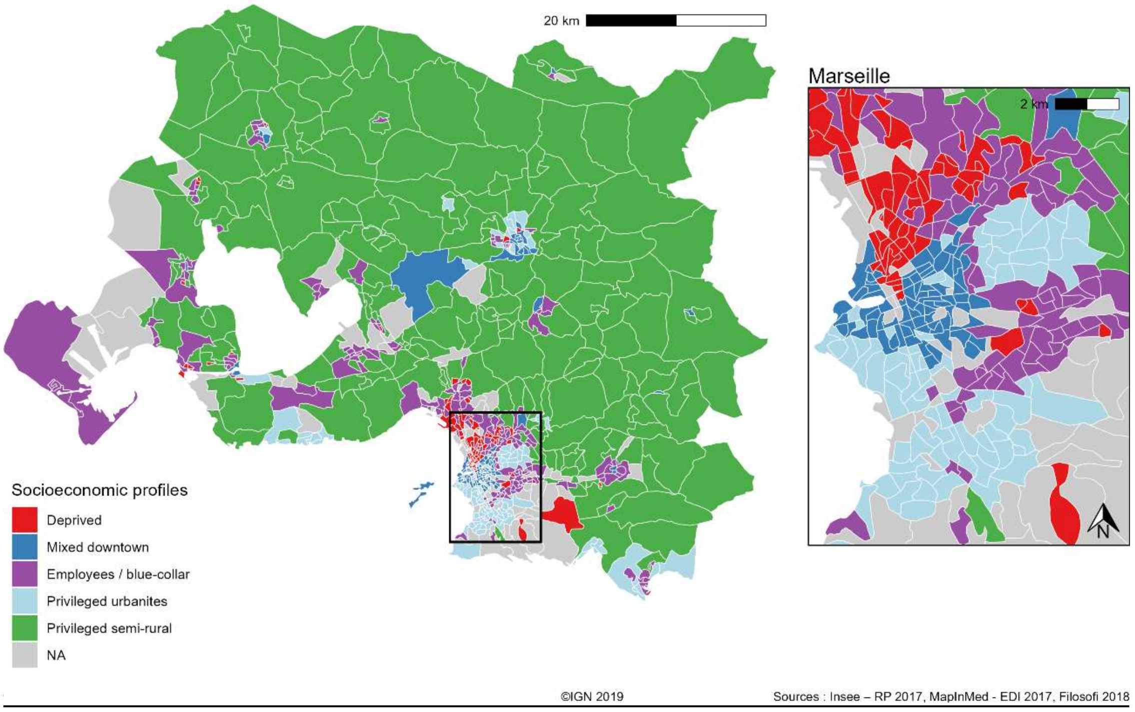


*Figure S1: Spatial distribution of the 5 socio-economic profiles of Aix-Marseille (deprived (n=89), mixed downtown (n=109), Employees / blue collar (n=190), privileged urbanites (n=108), privileged semi-rural (n=208))*

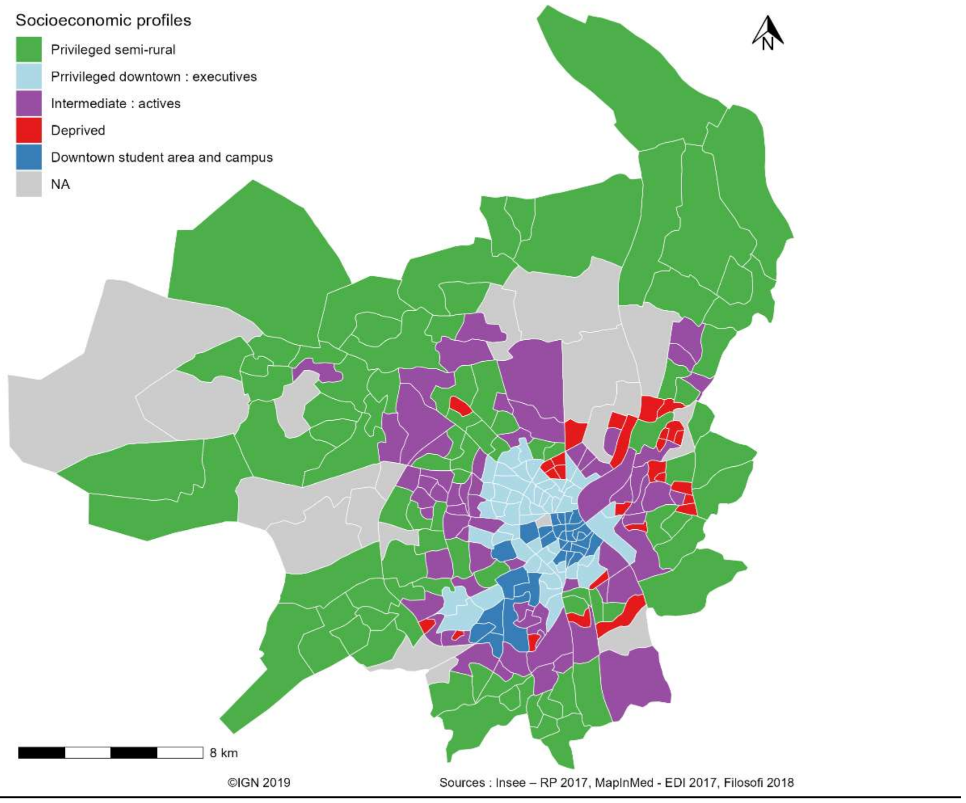


*Figure S2: Spatial distribution of the 5 socio-economic profiles of Bordeaux (privileged semi-rural (n=87), privileged downtown : executives (n=52), intermediate : actives (n=65), deprived (n=27), mi downtown (n=109), downtown student area and campus (n=28))*

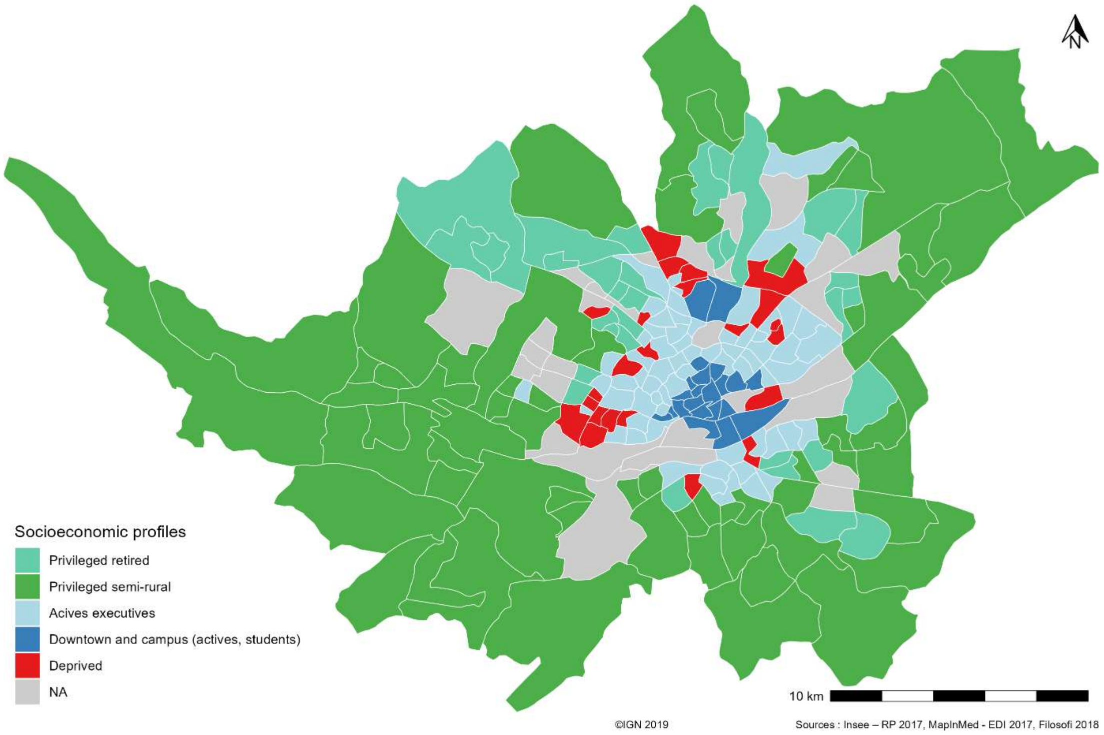


*Figure S3: Spatial distribution of the 5 socio-economic profiles of Nantes (privileged retired (n=33), privileged semi-rural : (n=62), intermediate : actives (n=65), actives executives (n=65), downtown and campus (actives, students) (n=22), deprived (n=23))*

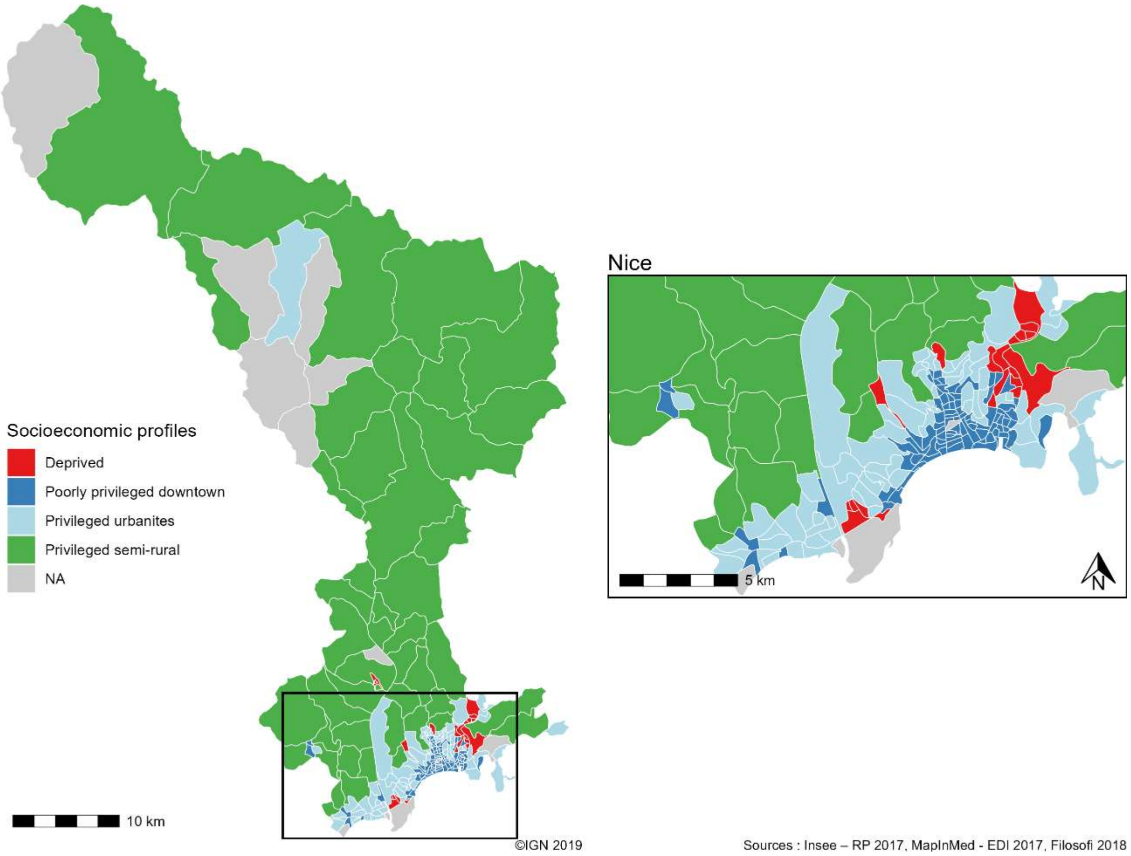


*Figure S4: Spatial distribution of the 4 socio-economic profiles of Nice (deprived (n=22), poorly privileged downtown (n=85), privileged urbanites (n=70), privileged semi-rural (n=47))*

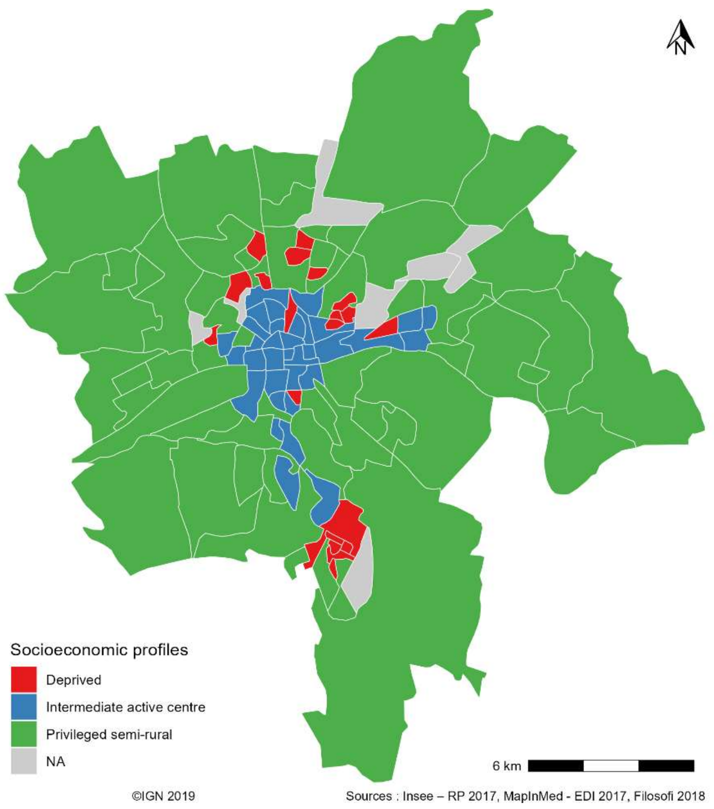


*Figure S5: Spatial distribution of the 3 socio-economic profiles of Orleans (deprived (n=20), intermediate active centre (n=33), privileged semi-rural (n=57))*

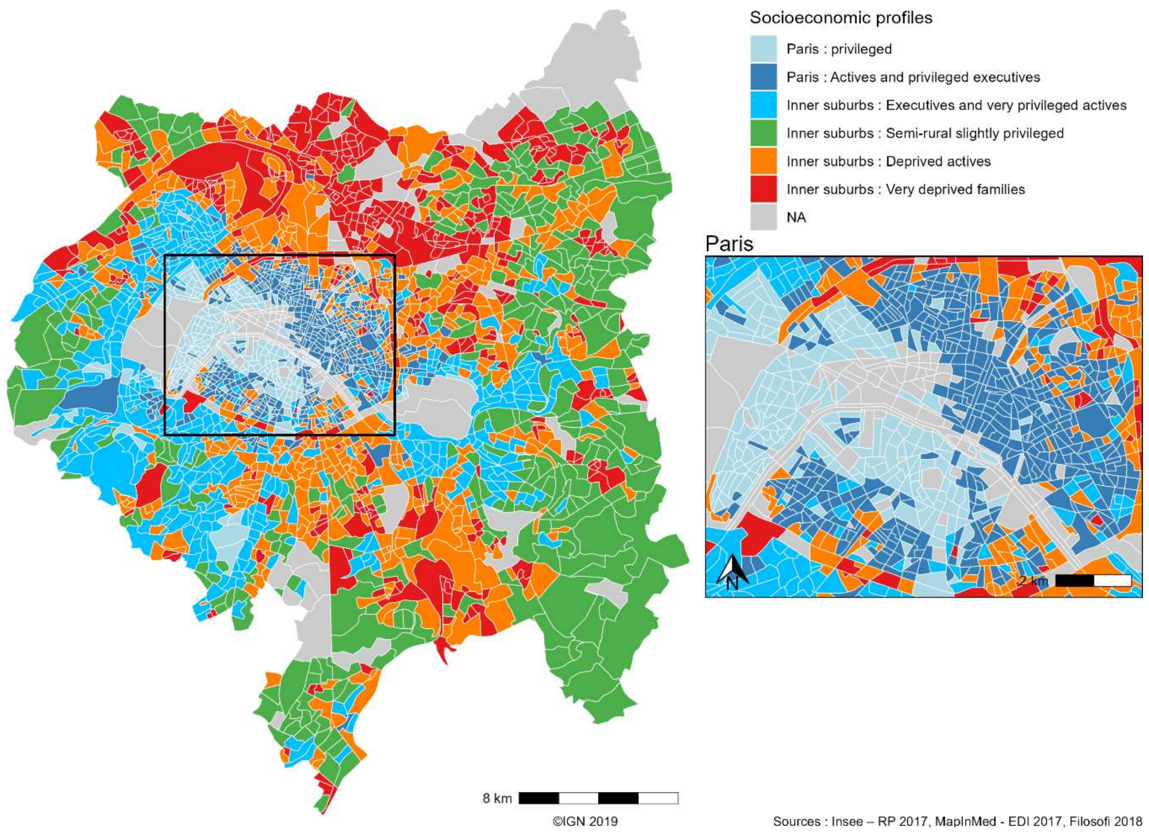


*Figure S6: Spatial distribution of the 6 socio-economic profiles of Grand Paris (Paris : privileged (n=223), Paris : actives and privileged executives (n=548), Inner suburbs : executives and very privileged actives (n=497), Inner suburbs : semi-rural slightly privileged (n=334), Inner suburbs : Deprived actives (n=601), Inner suburbs : very deprived families (n=469))*

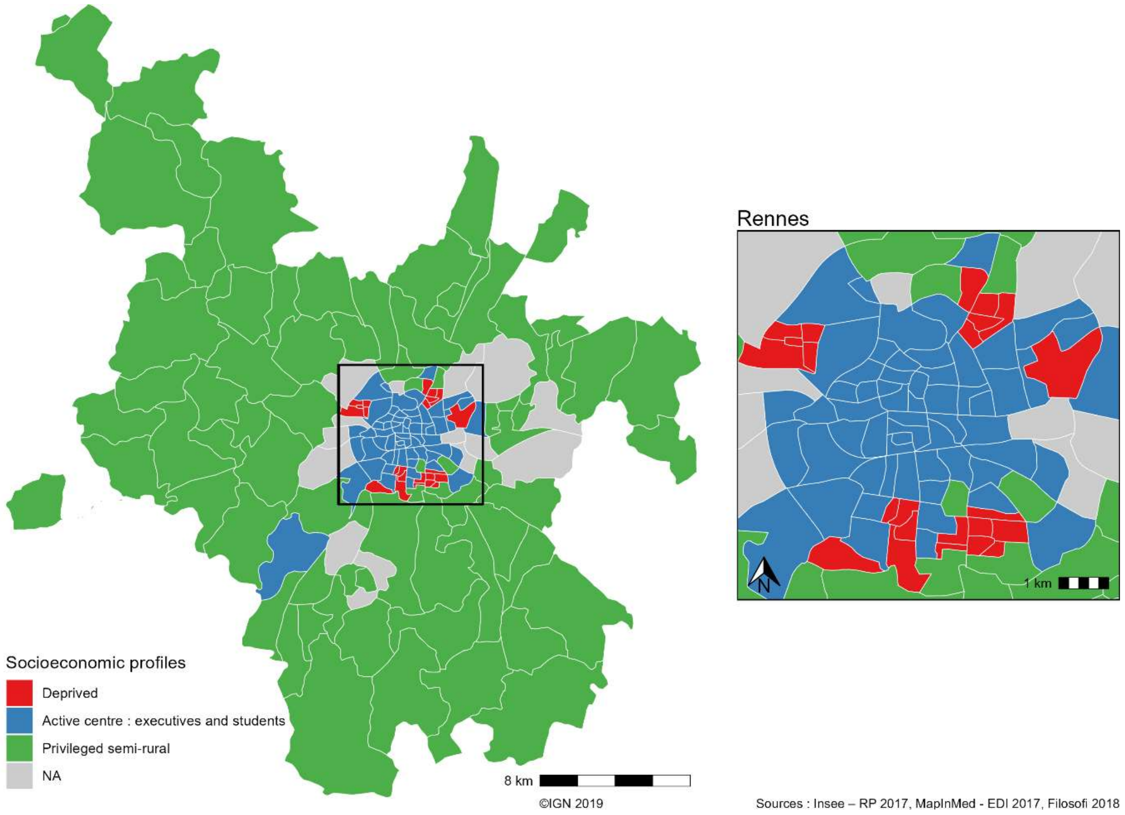


*Figure S7: Spatial distribution of the 3 socio-economic profiles of Rennes (deprived (n=23), active centre : executives and students (n=59), privileged semi-rural (n=76))*

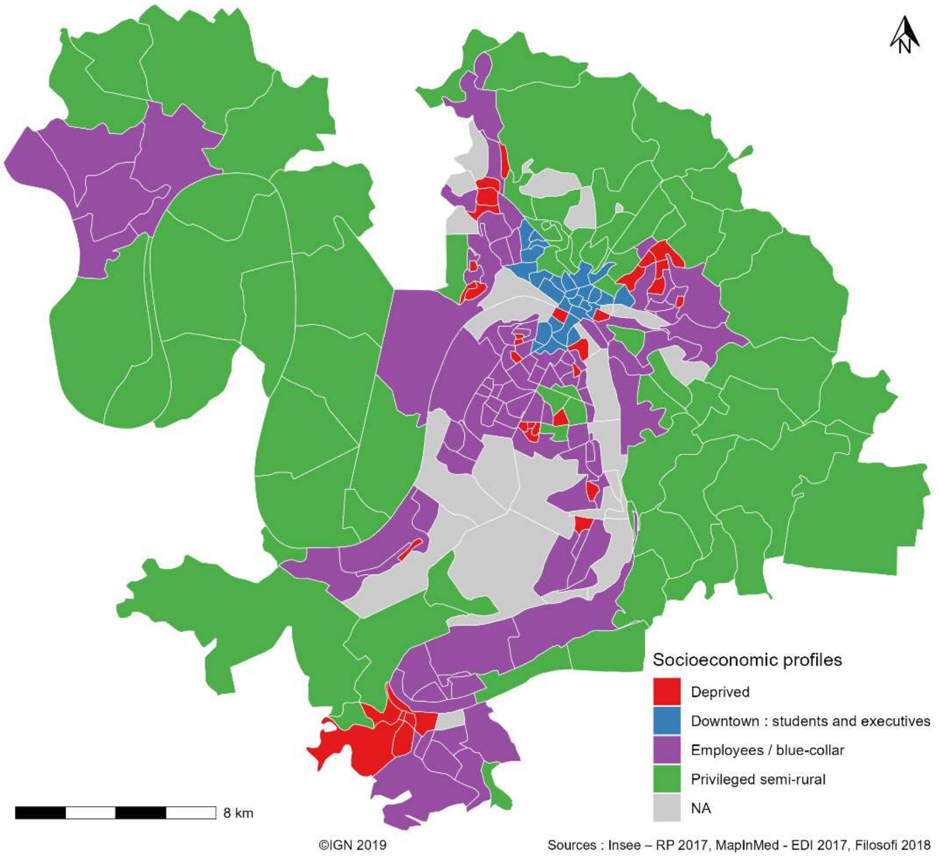


*Figure S8: Spatial distribution of the 4 socio-economic profiles of Rouen (deprived (n=34), downtown : students and executives (n=27), employees / blue-collar (n=89) , privileged semi-rural (n=67))*

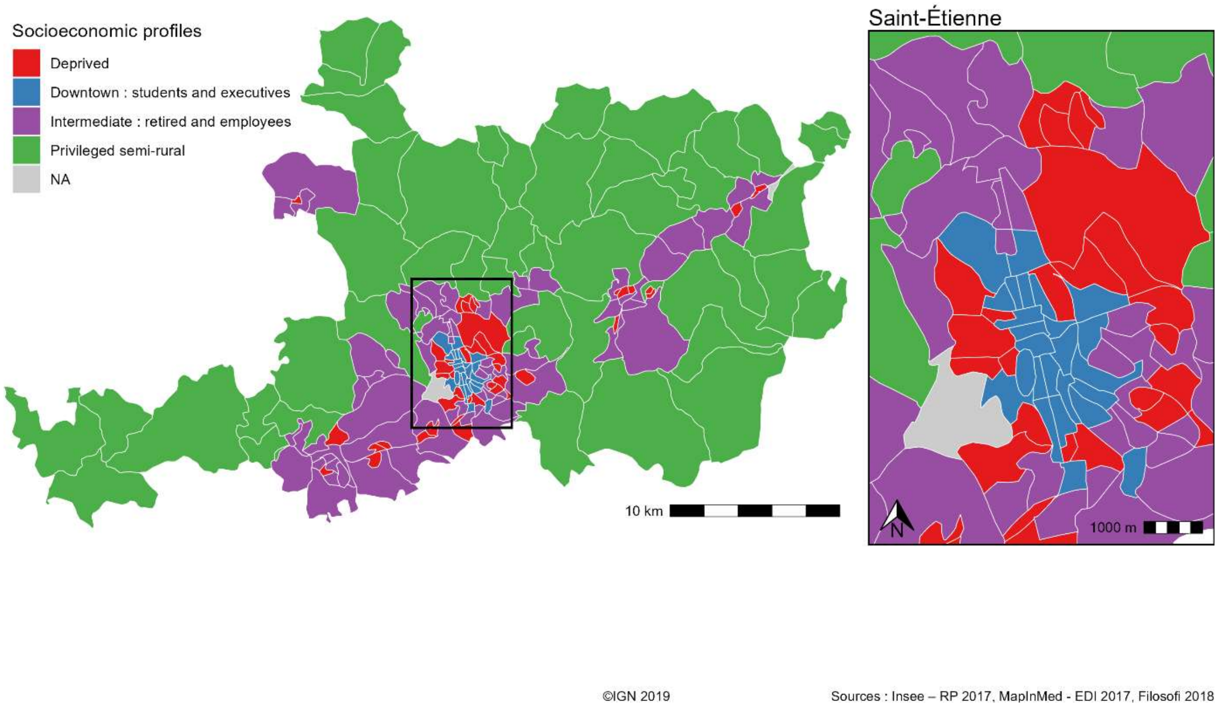


*Figure S9: Spatial distribution of the 4 socio-economic profiles of Saint-Etienne (deprived (n=43), downtown : students and executives (n=25), intermediate : retired and employees (n=69), privileged semi-rural (n=57))*

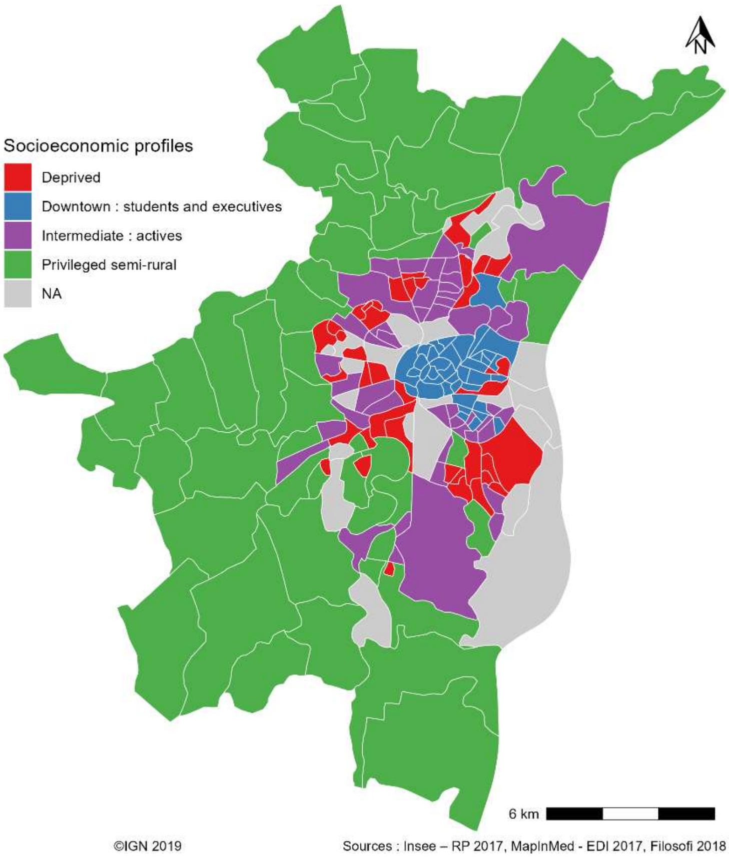


*Figure S10: Spatial distribution of the 4 socio-economic profiles of Strasbourg (deprived (n=40), downtown : students and executives (n=38), intermediate : actives (n=47), privileged semi-rural (n=49))*

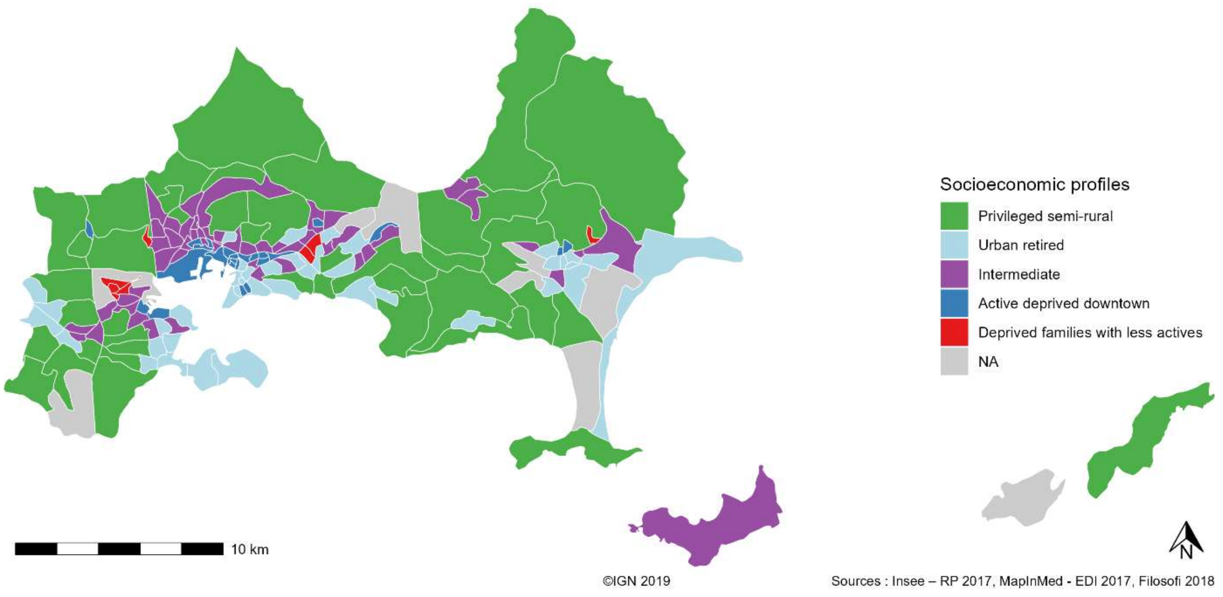


*Figure 11: Spatial distribution of the 5 socio-economic profiles of Toulon (privileged semi-rural (n=42), urban retired (n=41), intermediate (n=51), active deprived downtown (n=25) , deprived families with less activities (n=8))*

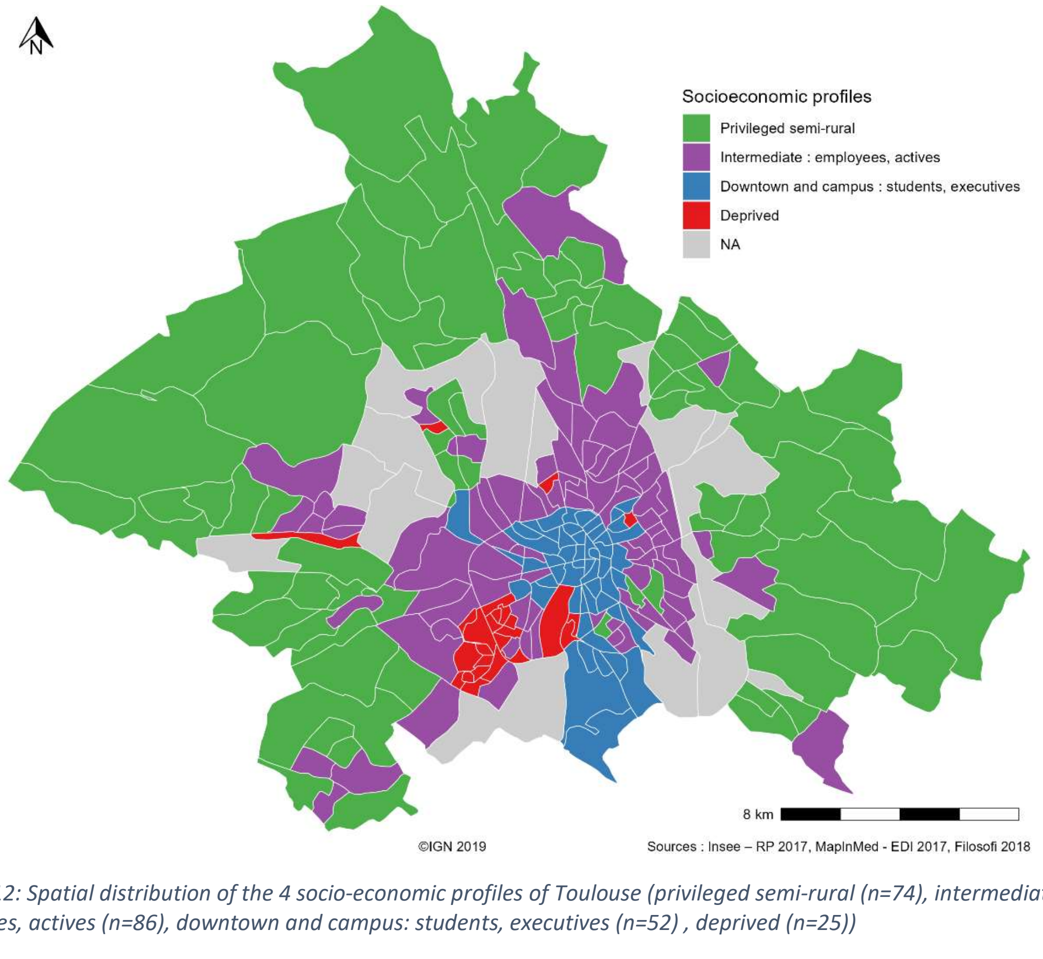


*Figure S12: Spatial distribution of the 4 socio-economic profiles of Toulouse (privileged semi-rural (n=74), intermediate: employees, actives (n=86), downtown and campus: students, executives (n=52) , deprived (n=25))*

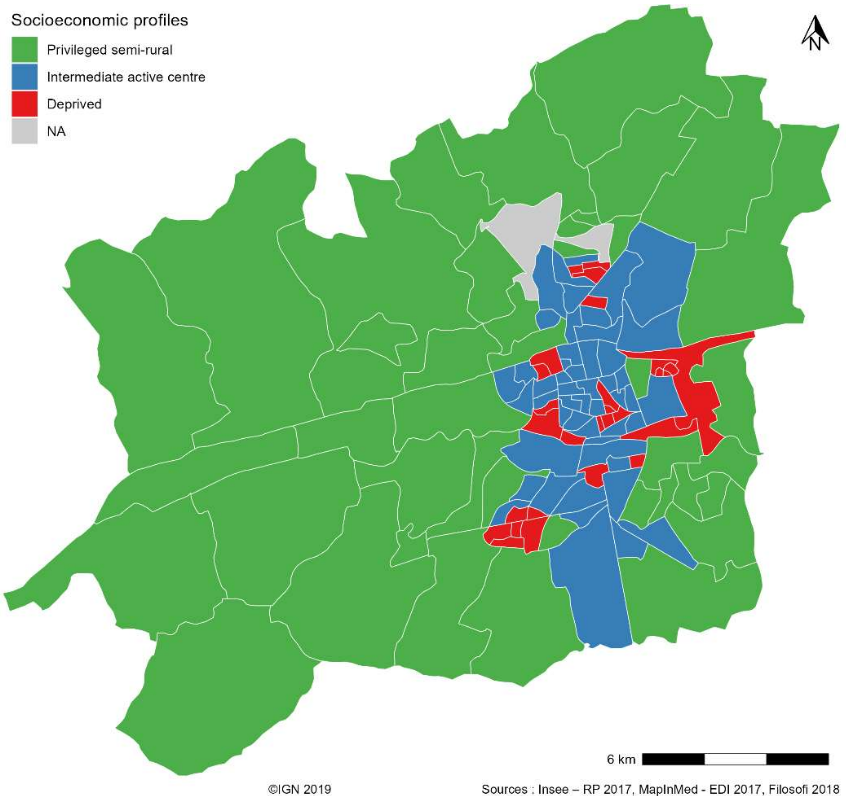


*Figure S13: Spatial distribution of the 3 socio-economic profiles of Tours (privileged semi-rural (n=50), intermediate active centre (n=41), deprived (n=27))*

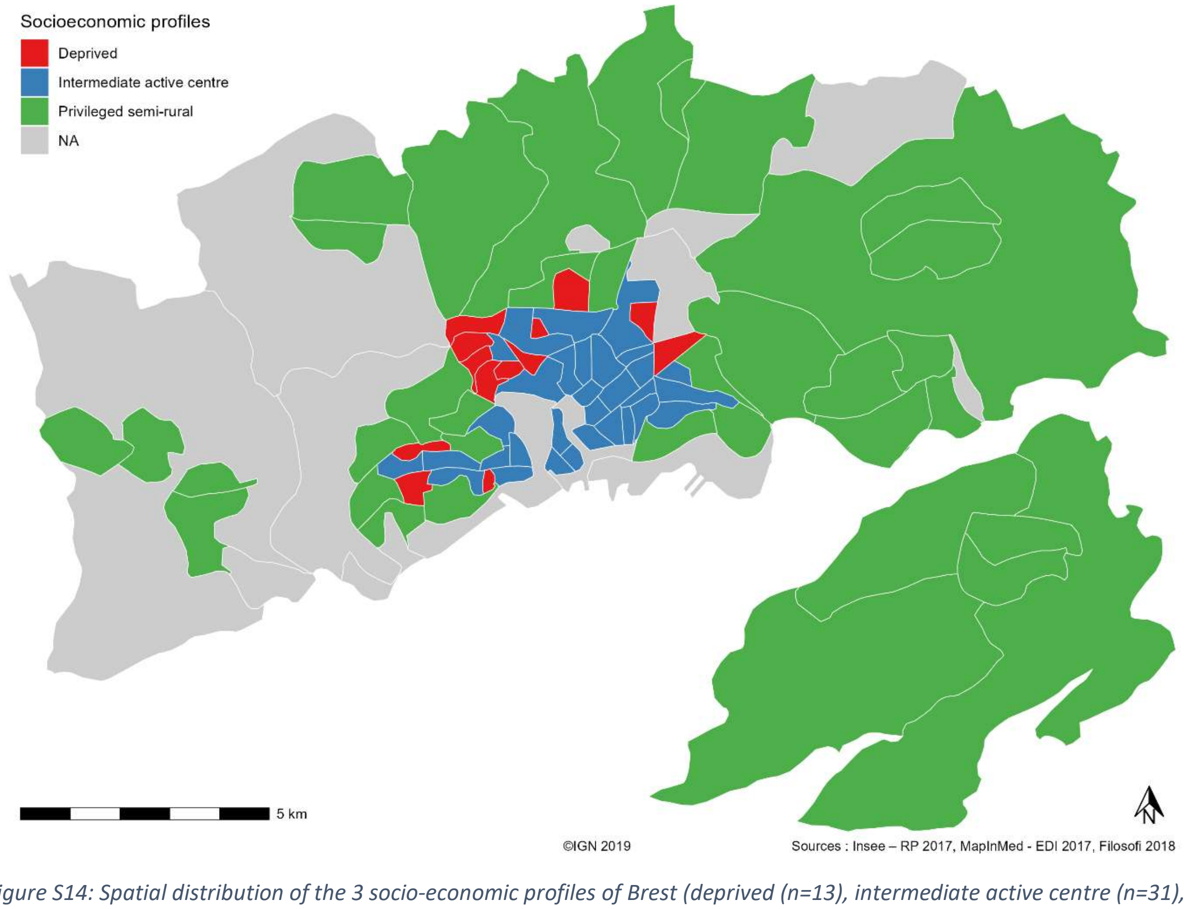


*Figure S14: Spatial distribution of the 3 socio-economic profiles of Brest (deprived (n=13), intermediate active centre (n=31), privileged semi-rural (n=36))*

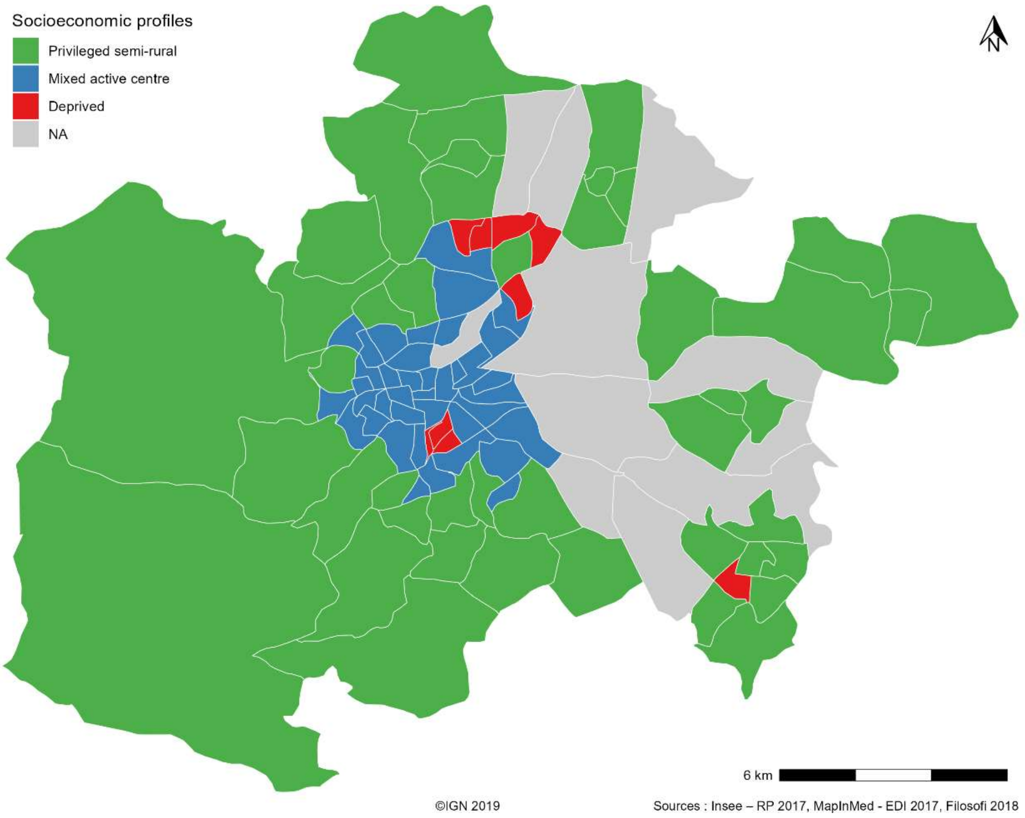


*Figure S15: Spatial distribution of the 3 socio-economic profiles of Clermont Auvergne (privileged semi-rural (n=44), mixed active centre (n=39), deprived (n=9))*

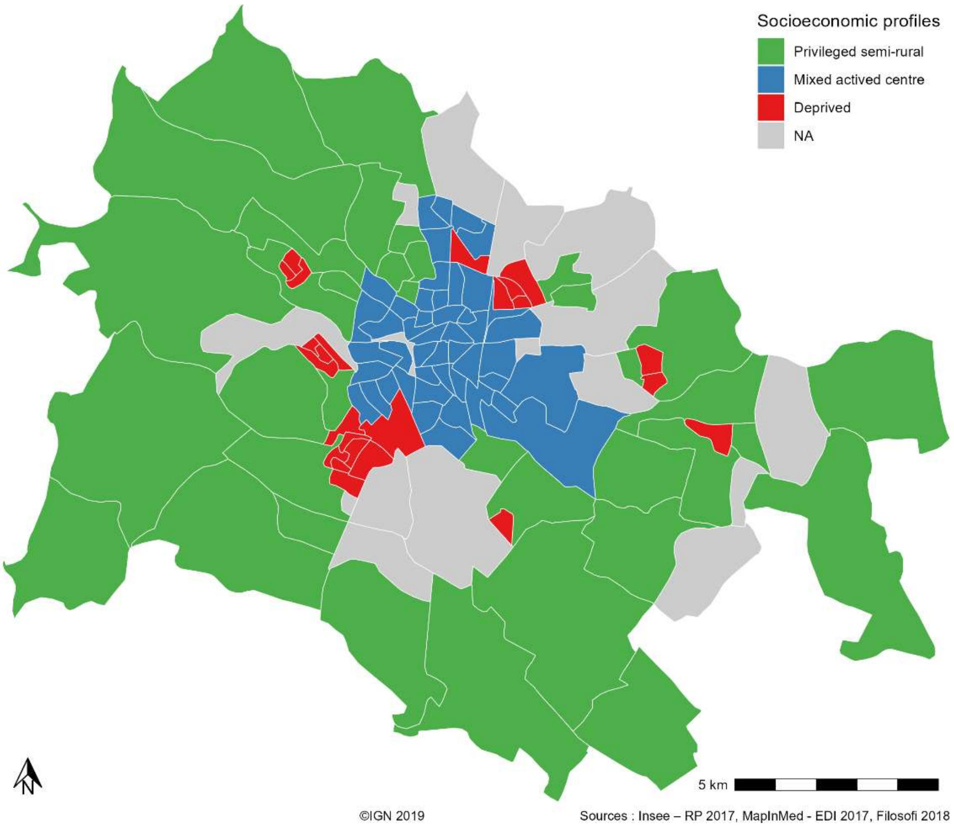


*Figure S16: Spatial distribution of the 3 socio-economic profiles of Dijon (privileged semi-rural (n=36), mixed active centre (n=47), deprived (n=24))*

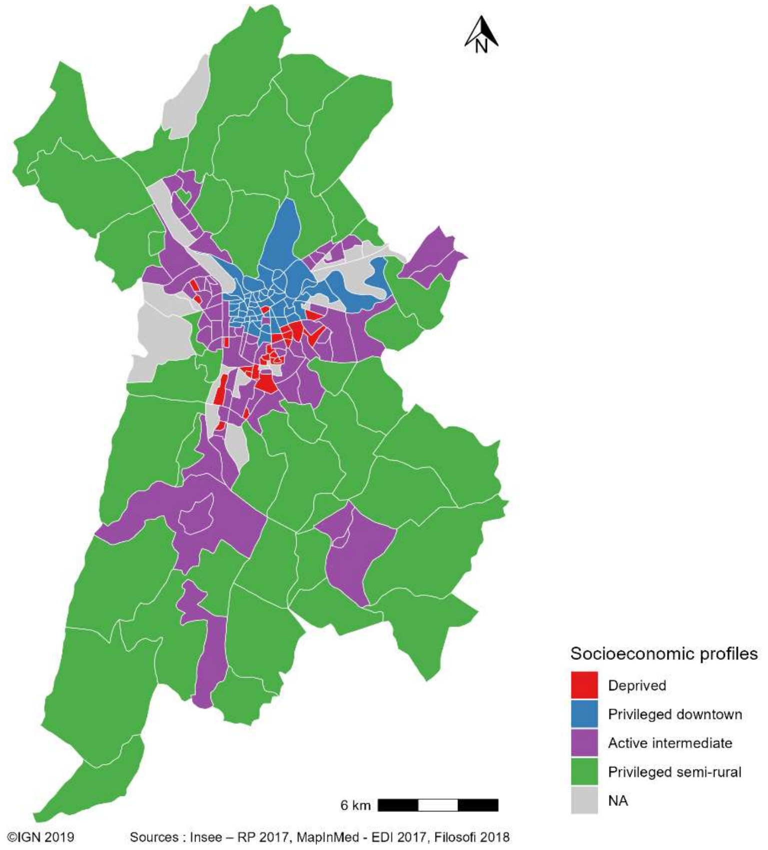


*Figure S17: Spatial distribution of the 4 socio-economic profiles of Grenoble (deprived (n=24), privileged downtown (n=50), active intermediate (n=67), privileged semi-rural (n=42))*

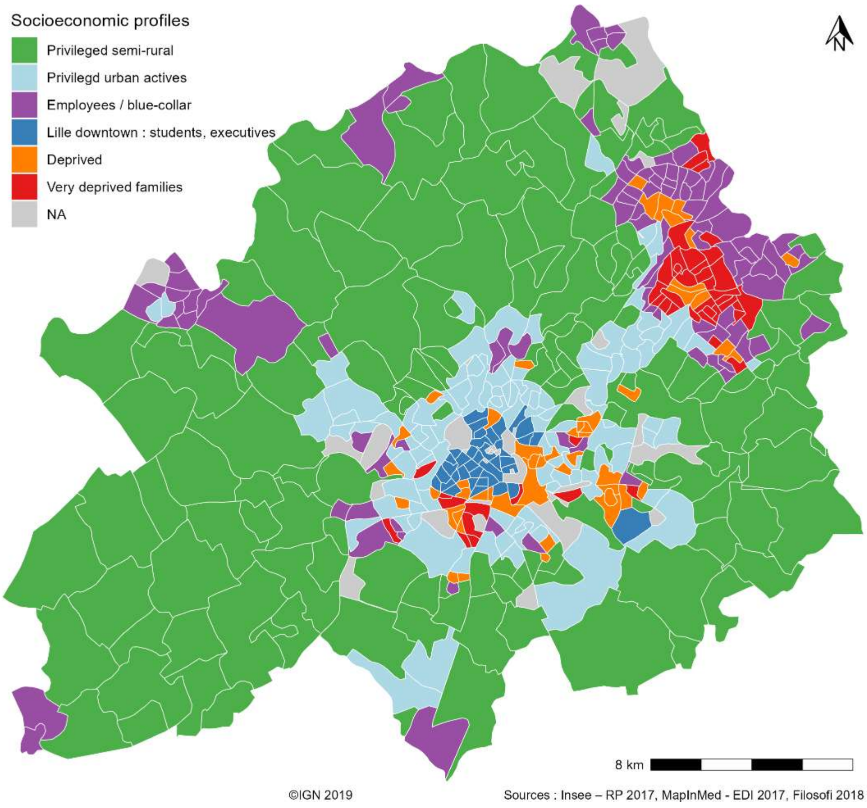


*Figure S18: Spatial distribution of the 6 socio-economic profiles of Lille (privileged semi-rural (n=144), privileged urban actives (n=104), employees / blue-collar (n=96), Lille downtown : students, executives (n=37), deprived (n=50), very deprived families (n=45))*

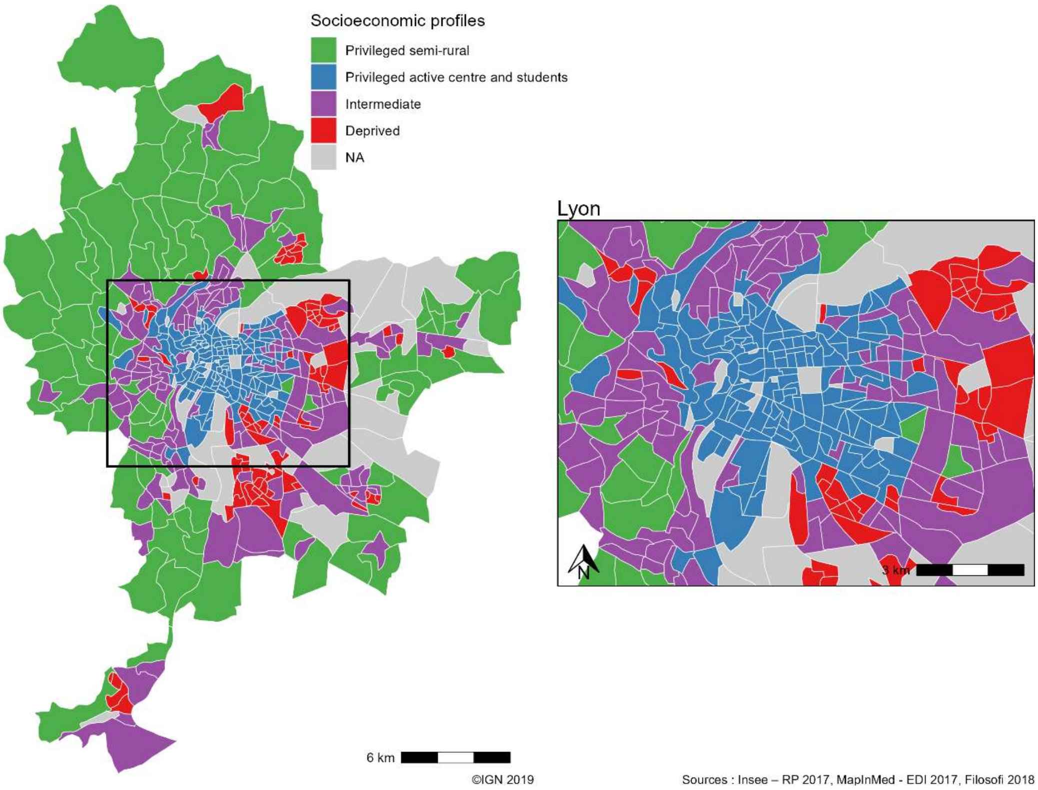


*Figure S19: Spatial distribution of the 4 socio-economic profiles of Lyon (privileged semi-rural (n=86), privileged active centre and students (n=149), intermediate (n=148), deprived (n=84))*

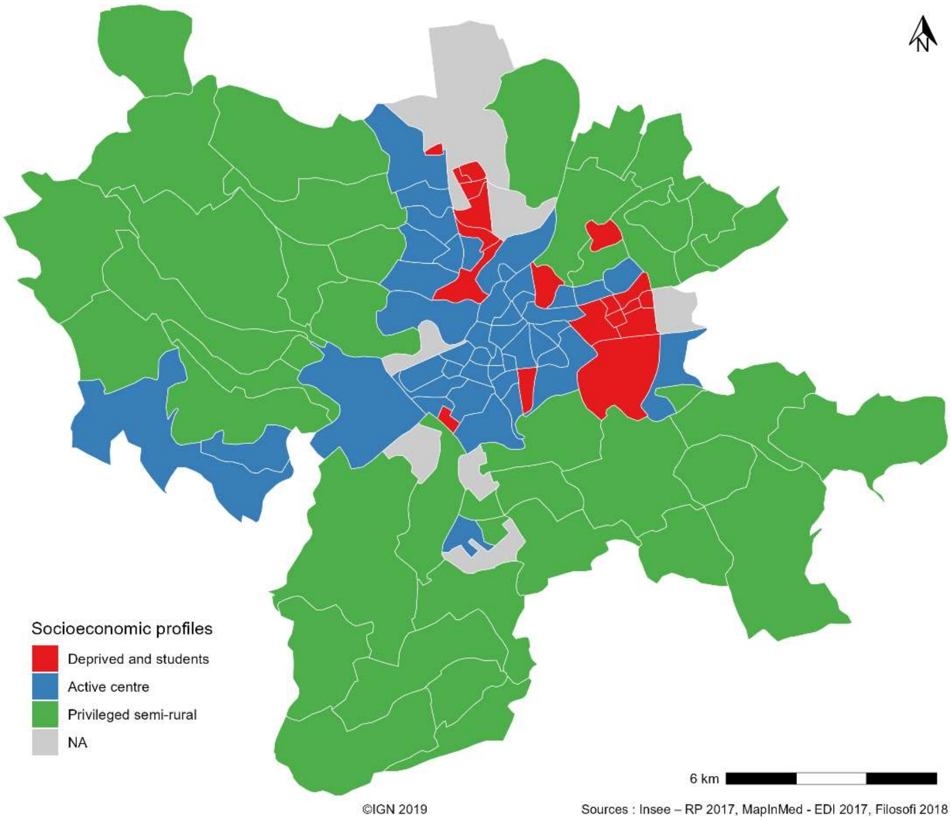


*Figure S20: Spatial distribution of the 3 socio-economic profiles of Metz (deprived and students (n=19), active centre (n=44), privileged semi-rural (n=43))*

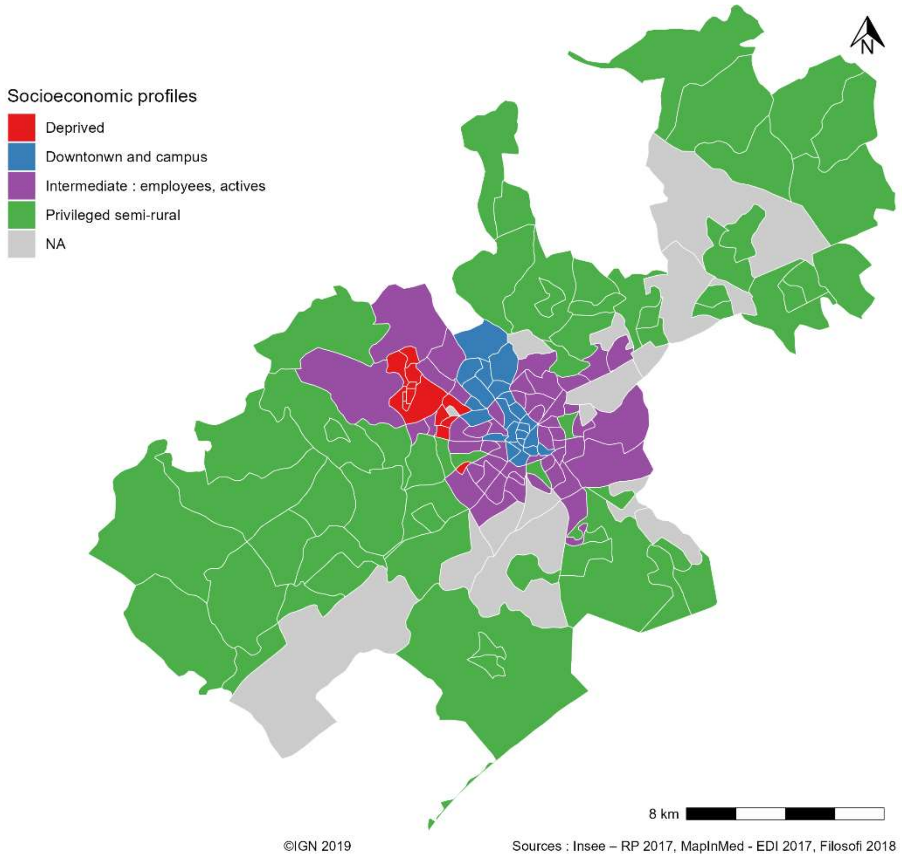


*Figure S21: Spatial distribution of the 4 socio-economic profiles of Montpellier (deprived (n=13), downtown and campus (n=22), intermediate : employees, actives (n=49), privileged semi-rural (n=60))*

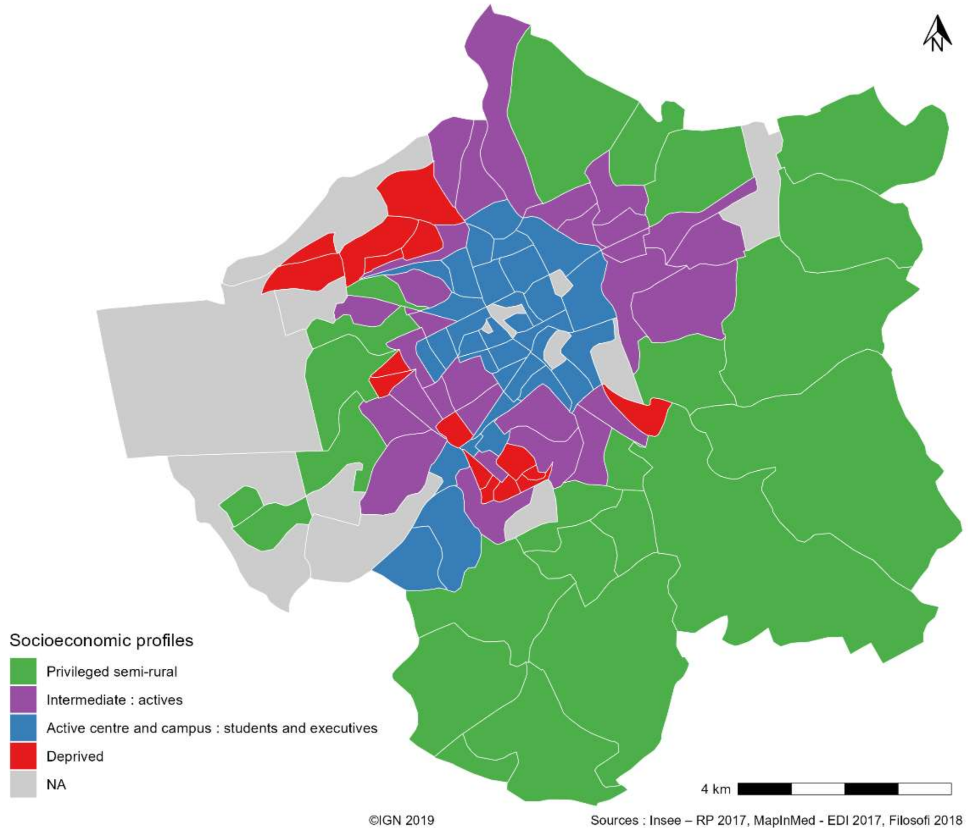


*Figure S22: Spatial distribution of the 4 socio-economic profiles of Grand Nancy (privileged semi-rural (n=24), intermediate: actives (n=29), active centre and campus: students and executives (n=33), deprived (n=16)*

#### Adjustment variables

##### Age distribution indicator

To create an indicator of the distribution of age, we selected 10 variables from the population census showing the percentage of inhabitants by age class (0–2,3-5,6-10,11-17,18-24,25-39,40-54,55-64,65-79,> 79). We calculed a Pearson assymetry coefficient with this formula [1] :

$$\frac{3 * (mean\ age - \ median\ age)}{standard\ deviation\ age}$$

We estimated the mean, median and standard deviation by taking the centre of the ten age classes. The Pearson asymmetry coefficient for age was interpreted as follows: a coefficient closer to 0 indicated a more symmetric age distribution. A positive coefficient suggested that the IRIS population tended toward younger ages, while a negative coefficient showed that the IRIS population tended toward older ages.

##### Population density

The population density was estimated by using the map of IRIS and a grid map that gave us residential areas where households lived. We calculated the area for each IRIS covered by the tiles, then we divided the total population of IRIS by this area and their base map of grid data to estimate the population density by IRIS.

##### Access to healthcare variables

Variables on access to healthcare come from the database equipment of INSEE and the Research, Studies, Evaluation and Statistics Directorate (DREES). We considered variables for the accessibility of testing. For each IRIS, we calculated several distances to the nearest medical laboratory and pharmacy where a COVID-19 test could be performed. We also estimated the number of pharmacies and medical laboratories per IRIS. We had other indicators describing the access to healthcare in general as the number of primary healthcare per IRIS. We used the localised potential accessibility (LPA), which is estimated at the municipal level and for each person, the number of consultations per year, taking into account the distance and the time to travel to the nearest medical doctor. Finally, we had the number of retirement homes per IRIS.

First, we chose the LPA because, unlike the other variables, it takes into account the time travel to consult a medical doctor. Secondly, we kept the number of retirement homes because the COVID-19 indicators included the results of these places. To have another variable in the IRIS scale on the accessibility of tests, we calculated the Spearman correlation and looked at the indicators that were least correlated with the LPA, the number of retirement homes and the deprivation score (EDI). Using this method, we selected the number of pharmacies and medical laboratories per IRIS.

##### Environment profiles

The raster data of land cover available on Copernicus contained 44 variables (31). Firstly, we grouped variables by following the Corine Land Cover nomenclature (level 2) [2]. Secondly, we made an additional grouping of variables to distinguish the artificial surfaces, the urban fabric and the other artificial surfaces as industrial, commercial, transport units, mine dumps and construction sites. We also grouped agriculture areas, forests, semi-natural areas and wetlands. Finally, we deleted water bodies because of IRIS border rivers which could bias analyses. To summarise, for each IRIS, we constructed 3 variables indicating the percentage of the urban fabric, other artificial surfaces (industrial, commercial…) and non-artificial surfaces covering the IRIS. We built environment profiles with the same method as socioeconomic profiles. For each metropolitan area, we injected the 3 environmental variables into a PCA, then we performed an HAC on axes with at least 99% inertia.

#### Univariate analysis

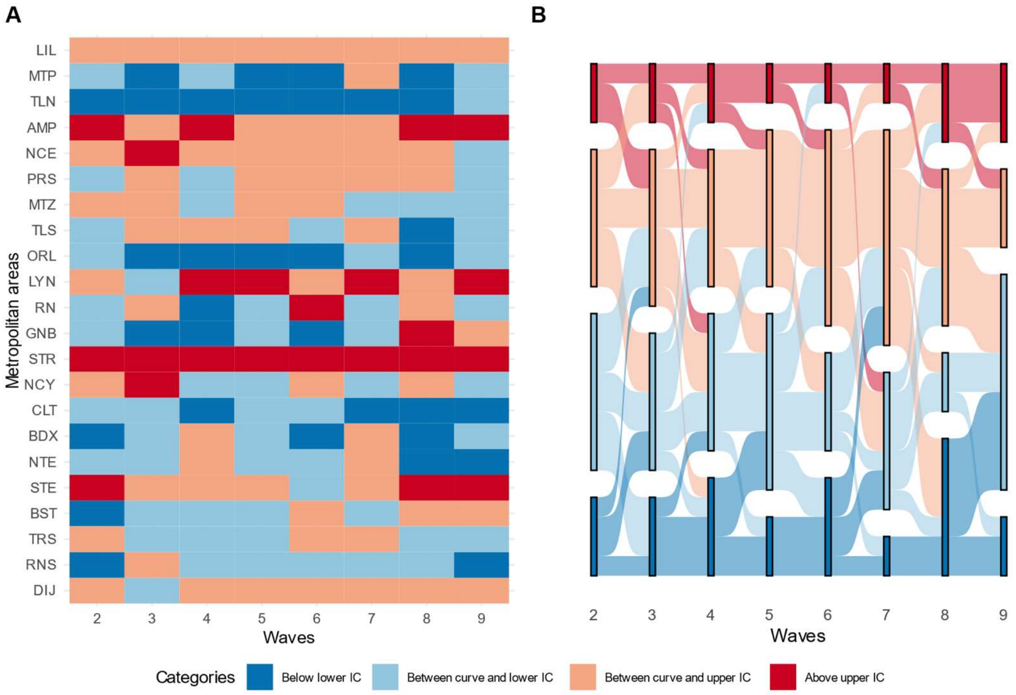


*Figure S23 :* Heatmap (A) and Sankey diagram (B) showing the evolution by metropolitan area and wave of the categories of testing rates, as a function of the median EDI of the most deprived profiles in the metropolitan areas.

The categories were defined based on their position relative to the adjustment curve derived from the LOESS regression and their confidence intervals (95% CI). On the heatmap (A), the metropolitan areas were ranked on the y-axis in ascending order of median EDI (Dijon had the lowest median EDI and Lille the highest).

*AMP = Aix-Marseille-Provence, BDX = Bordeaux, BST = Brest, CLT = Clermont Auvergne, DIJ = Dijon, GNB = Grenobles-Alpes, LIL = Lille, LYN = Lyon, MTP : Montpellier Méditerranée, MTZ = Metz, NCE = Nice Côte d'Azur, NCY = Grand Nancy, NTE = Nantes, ORL = Orléans, PRS = Grand Paris, RN = Rouen, RNS = Rennes, STE = Saint-Etienne, STR = Strasbourg, TLN = Toulon, TRS = Tours*

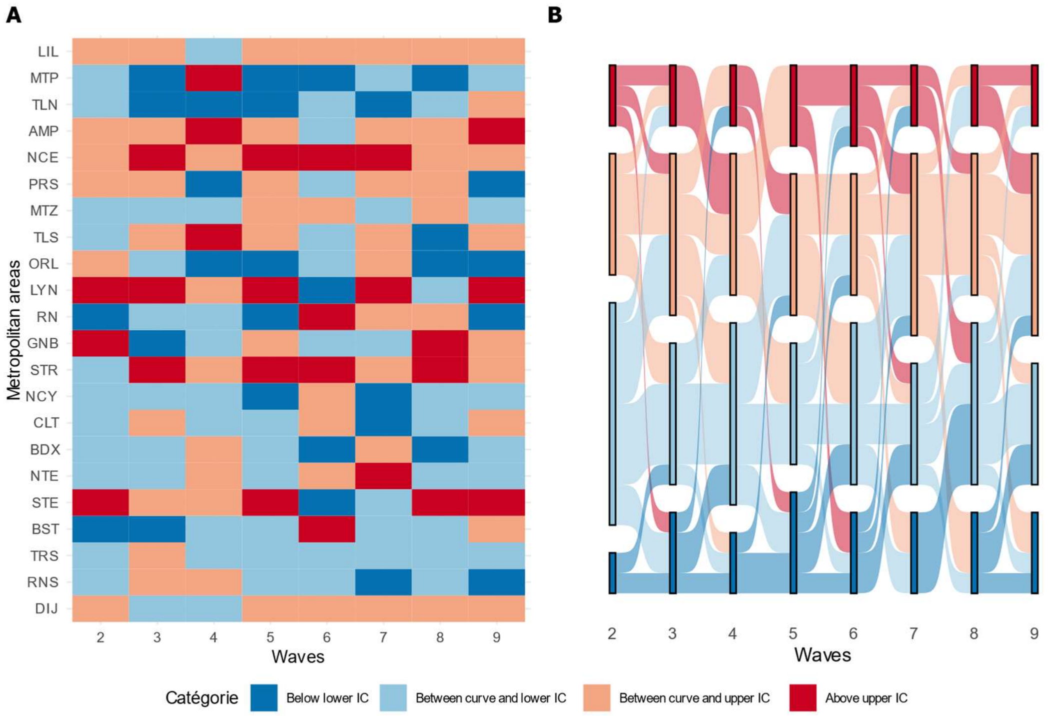


*Figure S24: Heatmap (A) and Sankey diagram (B) showing the evolution by metropolitan area and wave of the categories of incidence rates, as a function of the median EDI of the most deprived profiles in the 22 French metropolitan areas.*

*The categories were defined based on their position relative to the adjustment curve derived from the LOESS regression and their confidence intervals (95% CI). On the heatmap, the metropolitan areas were ranked on the y-axis in ascending order of median EDI (Dijon had the lowest median EDI and Lille the highest).*

*AMP = Aix-Marseille-Provence, BDX = Bordeaux, BST = Brest, CLT = Clermont Auvergne, DIJ = Dijon, GNB = Grenobles-Alpes, LIL = Lille, LYN = Lyon, MTP : Montpellier Méditerranée, MTZ = Metz, NCE = Nice Côte d'Azur, NCY = Grand Nancy, NTE = Nantes, ORL = Orléans, PRS = Grand Paris, RN = Rouen, RNS = Rennes, STE = Saint-Etienne, STR = Strasbourg, TLN = Toulon, TRS = Tours*

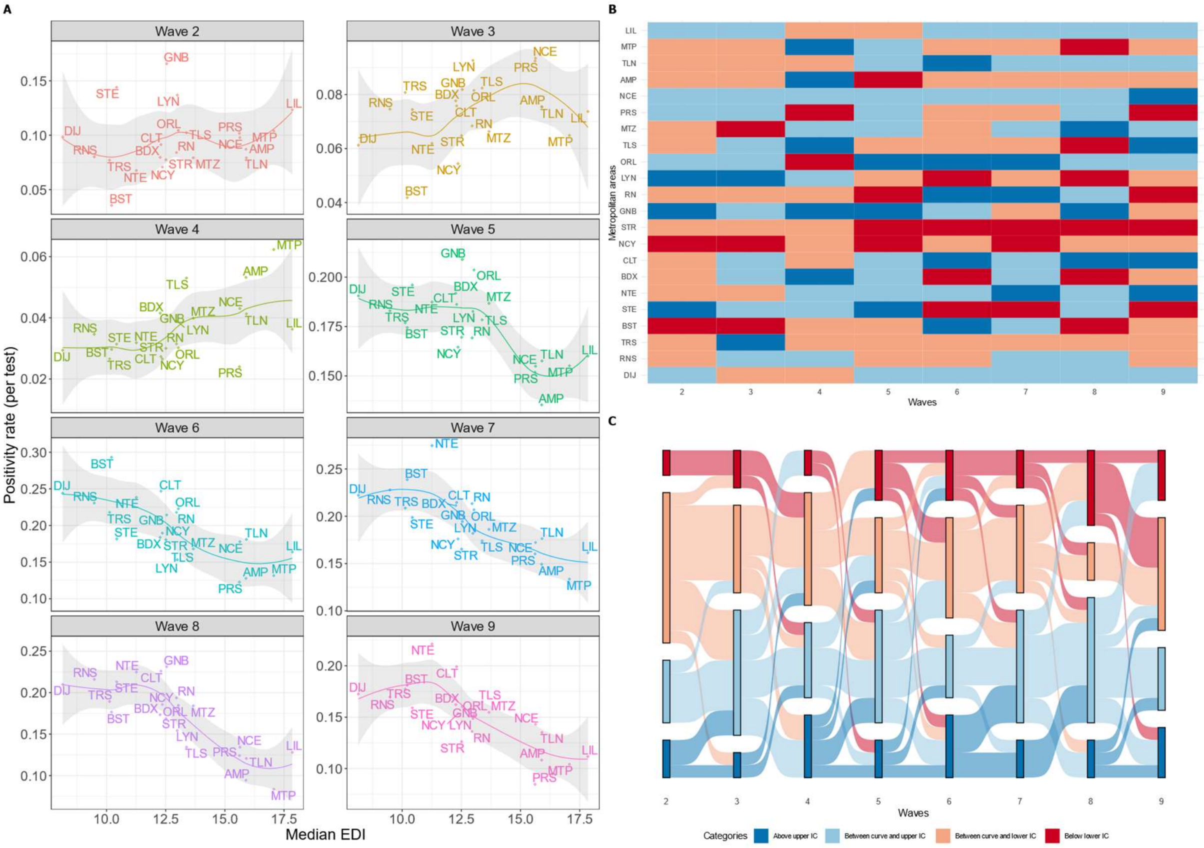


*Figure S25: Evolution by wave (waves 2 to 9) of positivity rates compared with the median deprivation score for the most deprived IRIS in the 22 French metropolitan areas (A), France, July 2020 – March 2023, n=1096.*

*AMP = Aix-Marseille-Provence, BDX = Bordeaux, BST = Brest, CLT = Clermont Auvergne, DIJ = Dijon, GNB = Grenobles-Alpes, LIL = Lille, LYN = Lyon, MTP : Montpellier Méditerranée, MTZ = Metz, NCE = Nice Côte d'Azur, NCY = Grand Nancy, NTE = Nantes, ORL = Orléans, PRS = Grand Paris, RN = Rouen, RNS = Rennes, STE = Saint-Etienne, STR = Strasbourg, TLN = Toulon, TRS = Tours*

*The graphs illustrate the smoothing curves (LOESS regressions with 95% confidence intervals colored in grey). Heatmap (B) and Sankey diagram (C) showing the evolution by metropolitan area and wave of the categories of incidence rates, as a function of the median EDI of the most deprived profiles in the metropolitan areas. The categories were defined based on their position relative to the adjustment curve derived from the LOESS regression and their confidence intervals (95% CI). On the heatmap, the metropolitan areas were ranked on the y-axis in ascending order of median EDI (Dijon had the lowest median EDI and Lille the highest).*

### Models

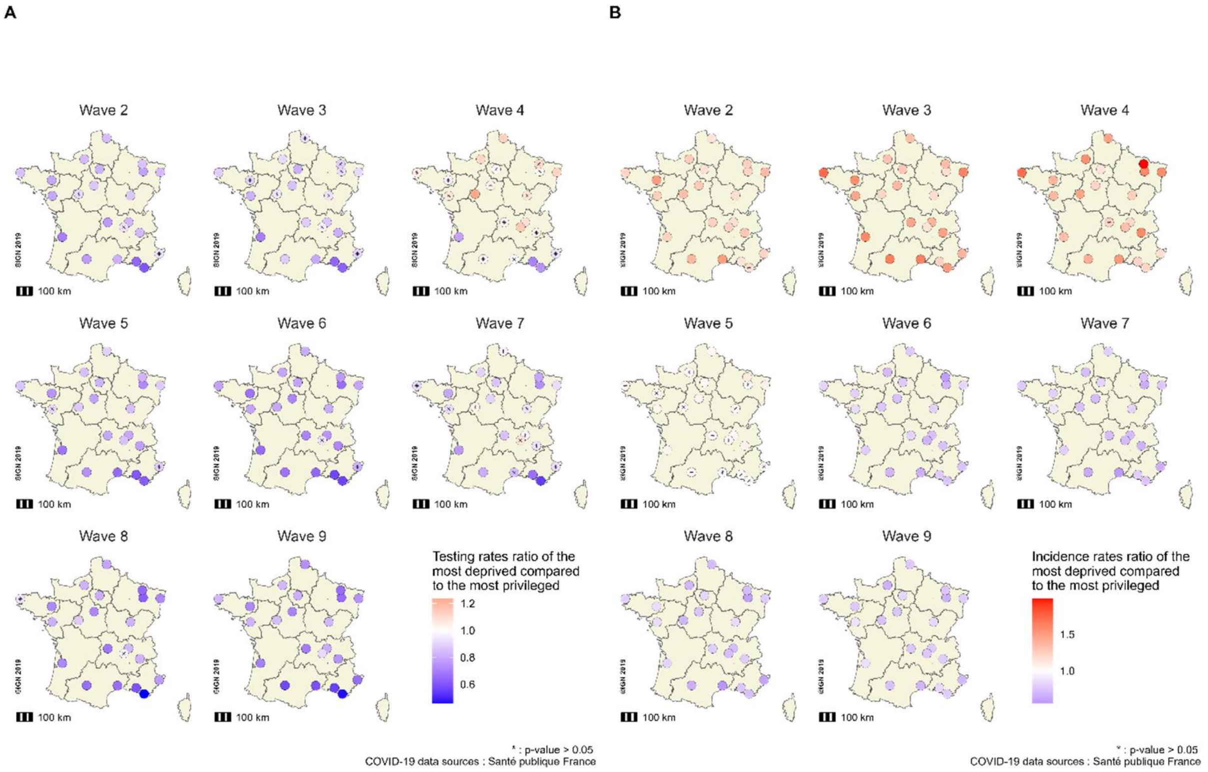


*Figure S26: Spatial distribution of testing (A) and incidence (B) rate ratios between the most deprived and most privileged profiles in the 22 French metropolitan areas, by wave (waves 2 to 9).*
*Ratios were computed from univariate (including random effects and spatial autocorrelation) GAM models per wave comparing testing and incidence rates for each metropolitan area between the socioeconomic profile with the lowest average EDI and the other profiles (French metropolitan areas, July 2020 – March 2023, n=7186).*
** : p-value > 0.05*

*Tableau S3: Testing rate ratios between the most deprived and most privileged profiles in the French metropolitan area of Nice with and without the IRIS outlier, by wave (waves 2 to 9).*
*Ratios were computed from univariate and multivariate GAM models per wave, comparing testing rates for each metropolitan area between the socioeconomic profile with the lowest average EDI and the other profiles (Nice, July 2020 – March 2023, n=224).*

| Waves | With IRIS outlier in Nice | | | | | | Without IRIS outlier in Nice | | | | | |
|---|---|---|---|---|---|---|---|---|---|---|---|---|
| | Univariate (with spatial autocorrelation and random effects) | | | Multivariate | | | Univariate (with spatial autocorrelation and random effects) | | | Multivariate | | |
| | TRR | IC 95% | p-value | aTTR | IC 95% | p-value | TRR | IC 95% | p-value | aTRR | IC 95% | p-value |
| 2 | 0.91 | 0.8 1.04 | 0.16 | 0.98 | 0.83 1.16 | 0.81 | 0.92 | 0.81 1.06 | 0.26 | 0.99 | 0.84 1.18 | 0.93 |
| 3 | 0.91 | 0.76 1.08 | 0.26 | 1.11 | 0.89 1.37 | 0.35 | 0.89 | 0.75 1.07 | 0.21 | 1.08 | 0.87 1.35 | 0.49 |
| 4 | 0.99 | 0.83 1.18 | 0.93 | 1.25 | 1 1.55 | 0.05 | 0.93 | 0.78 1.11 | 0.41 | 1.14 | 0.92 1.42 | 0.23 |
| 5 | 0.98 | 0.8 1.19 | 0.81 | 1.23 | 0.96 1.59 | 0.11 | 0.88 | 0.72 1.07 | 0.21 | 1.09 | 0.85 1.4 | 0.49 |
| 6 | 1.05 | 0.83 1.32 | 0.70 | 1.43 | 1.05 1.93 | 0.02 | 0.82 | 0.65 1.02 | 0.08 | 1.09 | 0.82 1.45 | 0.57 |
| 7 | 1.25 | 0.99 1.57 | 0.06 | 1.59 | 1.18 2.13 | 0 | 0.92 | 0.74 1.15 | 0.48 | 1.18 | 0.9 1.54 | 0.24 |
| 8 | 1.07 | 0.84 1.36 | 0.59 | 1.45 | 1.08 1.94 | 0 | 0.76 | 0.61 0.96 | 0.02 | 1.12 | 0.85 1.49 | 0.41 |
| 9 | 0.96 | 0.75 1.22 | 0.72 | 1.08 | 0.8 1.45 | 0.61 | 0.66 | 0.53 0.82 | 0 | 0.77 | 0.58 1.02 | 0.06 |

### Meta-regressions

A

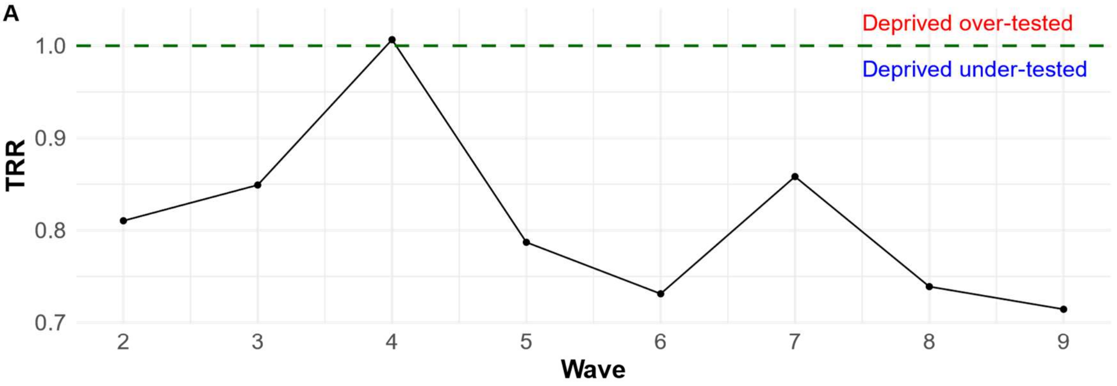


B

| Variable | Beta | 95% CI | p-value |
|---|---|---|---|
| Wave | -0.045 | [-0.062, -0.029] | <0.001 |
| Median cumulative vaccination rate (in %) | 0.0020 | [0.0008, 0.0033] | 0.018 |
| Median EDI of deprived profile | -0.020 | [-0.029, -0.011] | <0.001 |

*Figure S27: Mean of TRRs between the most deprived and most privileged profiles per epidemic wave (A) and results of meta-regression on logarithm of aTRRs in the 22 French metropolitan areas (B), July 2020 – March 2023 (n=176).*

*Green dotted lines corresponded to the testing equality point between the privileged and the deprived (TRR = 1). The table showed estimated betas for the three explanatory variables (wave, median cumulative vaccination rate and median EDI of deprived profile) with confidence intervals (95% CI) and p-values.*

A

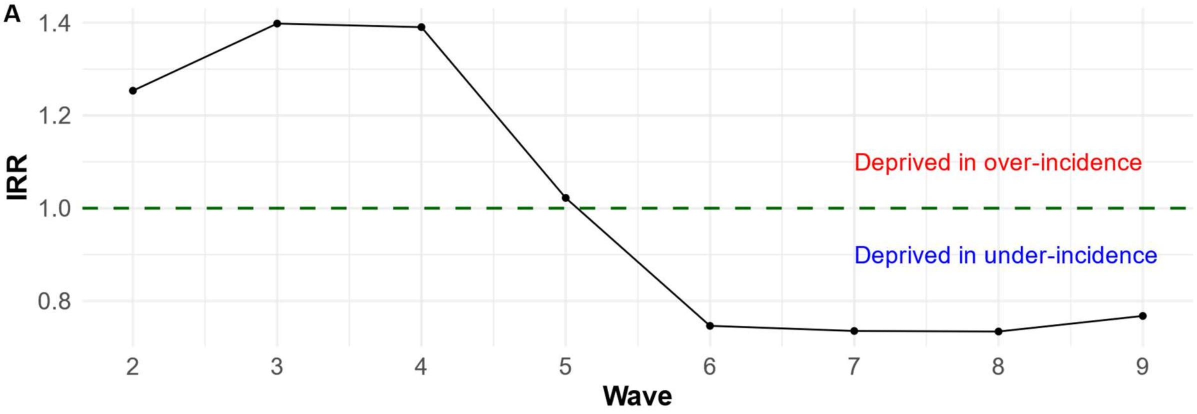


B

| Variable | Beta | 95% CI | p-value |
|---|---|---|---|
| Wave | -0.093 | [-0.112, -0.074] | <0.001 |
| Median cumulative vaccination rate (in %) | -0.0010 | [-0.0025, 0.0004] | 0.15 |
| Median EDI of deprived IRIS | -0.008 | [-0.018, 0.002] | 0.11 |

*Figure S28 : Mean of IRRs between the most deprived and most privileged profiles per epidemic wave (A) and results of meta-regression on logarithm of IRRs in the 22 French metropolitan areas (B), July 2020 – March 2023 (n=176).*

*Green dotted lines corresponded to green dotted lines corresponded to the testing equality point between the privileged and the deprived (IRR = 1). The table showed estimated betas for the three explanatory variables (wave, median cumulative vaccination rate and median EDI of deprived profile) with confidence intervals (95% CI) and p-values.*
